# Multi-institutional Normal Tissue Complication Probability (NTCP) Prediction Model for Mandibular Osteoradionecrosis: Results from the PREDMORN Study

**DOI:** 10.1101/2025.02.10.25322026

**Authors:** Laia Humbert-Vidan, Christian R. Hansen, Steven Petit, Carles Muñoz-Montplet, Abdallah S. R. Mohamed, Deborah P. Saunders, Vinod Patel, Gerda M. Verduijni, Wilma D. Heemsbergen, Arjen van der Schaaf, Max Witjes, Suzanne de Vette, Abdul A. Khan, Jordi Marruecos Querol, Irene Oliveras Cancio, Mike Oliver, Peter Reich, Stacey A. Santi, Andrew G. Pearce, Stephen Y Lai, Andrew P. King, Johannes A. Langendijk, Jørgen Johansen, Amy C Moreno, Clifton D. Fuller, Lisanne V. van Dijk, Teresa Guerrero Urbano

**Author notes:** Joint last authors (Lisanne V. van Dijk, Teresa Guerrero Urbano).

## Abstract

**Background:** Mandibular osteoradionecrosis (ORN) is a severe late complication affecting patients with head and neck cancer (HNC) treated with radiotherapy that significantly impacts patients’ quality of life and can require costly interventions. While radiation dose is a key factor, other clinical and demographic risk factors influence ORN development. Previous predictive models have primarily been single-institutional, limiting their generalizability. The PREDMORN Consortium was established to address these limitations. In this first analysis, we have aimed to reproduce existing statistical association and modelling analyses on the largest and most diverse mandibular ORN cohort worldwide to allow comparison with previous studies. As such, we have developed, tested and externally validated a multi-institutional normal tissue complication probability (NTCP) model for mandibular ORN.

**Methods:** This retrospective multi-institutional study included 1,184 HNC patients (389 ORN cases) from seven institutions. Clinical, demographic, and dosimetric (DVH) variables were analysed to develop a prediction model (any grade of ORN vs. no ORN) using forward stepwise logistic regression with correlation-based variable pre-selection. The ORN NTCP model was developed on 80% of data from six institutions, tested on the remaining unseen 20%, and externally validated on the seventh institution’s dataset.

**Results:** Key predictors of ORN were D30%, V70Gy, pre-RT dental extractions, and smoking status. The ORN NTCP model demonstrated good calibration and predictive performance, with AUCs of 0.69 for internal testing and external validation, which improved when tested on a sub-cohort of oropharyngeal and locally advanced larynx/hypopharynx cancer cases (AUCs of 0.75).

**Conclusion:** The PREDMORN NTCP model is the largest multi-institutional effort to predict ORN risk in HNC patients. We provide guidance on how to adjust the NTCP predicted probabilities for differences in target population baseline ORN risk to facilitate application of the model. Future research will focus on incorporating imaging-based spatial data and further external validation to enhance clinical applicability.

## 1. Introduction

Radiation therapy (RT), alone or in combination with chemotherapy and surgery, is the mainstay treatment for head and neck cancer (HNC). While current external beam RT delivery techniques allow for superior sparing of normal tissues in the head and neck region, treatment-related toxicities are still significant. Mandibular osteoradionecrosis (ORN) is a debilitating late side effect that develops in 4-15%^1^ of patients with HNC treated with RT. ORN has a detrimental impact on patients’ quality of life^2^, often necessitating costly clinical interventions^3^. When the mandible is irradiated, its vascularisation is compromised, resulting in hypocellularity of the bone and soft tissues^4,5^. Eventually, this can develop into necrosis of the bone, which can be diagnosed at different disease stages, ranging from subtle radiographic thinning of the cortical bone to exposed bone or even fracture of the mandible^6^. While radiation dose plays an obvious role, understanding the combined contribution of other associated factors for ORN is key to developing effective mitigation strategies. This is particularly important in the context of the observed rise in HPV-associated oropharyngeal cancer (OPC), which has led to a growing population of younger patients with longer overall survival and, therefore, a longer window during which ORN can develop.

While numerous studies^7–13^ have previously investigated the associations between ORN with demographic, clinical, and dosimetric factors, most models have been developed and validated using only single-institutional cohorts. Comparisons between studies is not straightforward due to the large diversity of cohorts, treatment techniques, ORN classification systems and inclusion criteria considered. Moreover, the clinical adoption of validated and generalizable conclusions is hampered by the limited size of these datasets, driven by the naturally low prevalence of ORN. For example, a normal tissue complication probability (NTCP) model for mandibular ORN was developed by van Dijk et al.^7^, using a large single-institution cohort, based on dose-volume metrics and clinical and demographic variables. While this model holds significant clinical value for the institution in which it was developed, its generalizability to other settings may be limited due to potential differences in treatment and patient characteristics.

To address these limitations, the PREDMORN (PREDiction models for Mandibular OsteoRadioNecrosis) Consortium14 was created, currently consisting of seven European and North American institutions: Guy’s and St Thomas’ NHS Foundation Trust (GSTT, United Kingdom), The University of Texas MD Anderson Cancer Center (MD Anderson, United States), Odense University Hospital (OUH, Denmark), Erasmus Medical Center (EMC, Netherlands), University Medical Center Groningen (UMCG, Netherlands), Catalan Institute of Oncology (ICO, Spain) and Health Sciences North Research Institute (HSNRI, Canada).

In this study, the PREDMORN Consortium aimed to i) curate the largest and most diverse ORN real-world dataset ever considered worldwide, ii) identify independent dosimetric and clinical predictors of ORN and iii) develop, test and externally validate an NTCP prediction model for ORN.

## 2. Materials and methods

### 2.1. Study design and patient selection

Following the required ethics approvals at each institution (GSTT: REC reference 18/NW/0297, IRAS project ID: 231443; MD Anderson: RCR-03-800; EMC: EMC17404; UMCG: NCT02435576; OUH: approval by The Danish Data Protection Agency [Jr. 16/29136] and The Danish Patient Safety Authority [Jr. 3-3013-1798/1/]; ICO: CEIC approval date 22/06/2015; HSNRI: REB number 23-038), subjects with mandibular ORN were retrospectively selected from their entire HNC population following the inclusion/exclusion criteria described in the study protocol^14^ (supplementary Table A1). The control subjects were selected following an approximate 2:1 matched control-case approach, as not all centres could retrieve data for their entire HNC population. As a result, the ORN incidence in this study does not represent the actual HNC population ORN rates which ranged from 5.1% to 13.7% across the different participating institutions. According to the study protocol^14^, matching was based on the primary tumour site and treatment year. The following primary tumour site groups were considered: oropharyngeal cancer (OPC), oral cavity cancer (OCC), larynx/hypopharynx cancer, and others, the latter including cases of salivary gland cancer, nasopharynx cancer, and unknown primary tumour sites.

### 2.2. Data

#### Clinical data

Clinical data was collected by each institution using a standardised data collection template with pre-defined variable codes (supplementary Table A2). Due to the retrospective study design, some centres encountered missing data. Variables with a percentage of missing data greater than 10% over the entire cohort were excluded from the analysis. Patients with missing values in the included variables were excluded. Based on the calculated percentages of missing data (supplementary tables B1 and B2), the following subset of clinical and demographic variables were selected for statistical analyses: sex (binary, male vs. female), age (continuous), mandible volume in cm^3^ (continuous), smoking status (binary, current vs. never/former), pre-RT dental extractions (binary, yes vs. no), chemotherapy (binary, yes vs. no), PORT (binary, PORT vs. primary RT), RT technique (binary, IMRT vs. VMAT), and fraction size (> vs. ≤ 2 Gy/fraction).

#### Dosimetric data

Segmentation of the mandible, including condyles and excluding teeth, was performed manually or using atlas-based auto-segmentation software (e.g., ADMIRE, Elekta, Sweden) on the treatment planning computed tomography images. Dosimetric data for the mandible structure were extracted from the treatment planning systems (TPS) as the cumulative DVH in absolute dose (Gy) and relative volume (%), and the following dosimetric variables were calculated: D_2%_ (where D_x%_ corresponds to the dose received by X% of the mandible volume), D_5%_-D_95%_ in 5% steps, D_98%_, V_5Gy_-V_70Gy_ (where V_XGy_ is the volume of the mandible that receives at least XGy) in 5 Gy steps. No EQD2 correction was performed; instead, the prescribed fraction size was considered as a variable to account for the potential effect of differences in fractionation schemes.

#### Clinical endpoint

Due to the variability between centres in ORN grading system (e.g., Notani^15^, Tsai^16^, see supplementary Table B3) and ORN intervention strategies, a uniform classification system across all participating centres could not be realised. The Notani ORN classification system was agreed upon for the study. While re-classification of cases using the Notani system was carried out retrospectively by some of the participating centres, direct translation from the Tsai system to the Notani system was not possible for other centres: Notani’s system is based on anatomical boundaries (ORN confined vs. beyond the alveolar bone) whereas the Tsai system grades the severity of the bone exposure based on the required clinical intervention. Therefore, the endpoint considered in this study was dichotomised as no ORN vs. any stage of ORN, as determined clinically and/or radiologically as per centre-specific protocols.

### 2.3. Statistical analyses and NTCP model development

Data from six institutions were used to train and test set the described model. Data from one institution, chosen without prior knowledge and in an unbiased manner, was kept as the independent dataset for external evaluation of the model. The model development dataset was partitioned into training (80%), and independent test (20%) sets using a stratified split (i.e., equal ratio of ORN vs. control subjects in both sets). Bootstrapping was performed to assess the robustness of variable selection and model performance. Finally, data from the seventh centre was used for external validation of the models.

For explorative purposes, univariable logistic regression analysis was performed on the full training set to assess the individual statistical relevance of both the clinical and dosimetric variables. Variables were ranked according to their level of significance (p-value) with regard to their association with ORN. For the NTCP model development, a multivariable forward stepwise logistic regression with a prior variable pre-selection approach was followed. First, redundant variables were excluded through a pre-selection procedure to avoid multicollinearity: if the correlation between pairs of variables was high (Spearman correlation coefficient > 0.8) then the variable of that pair with the lowest univariable association to ORN (lowest p-value) was omitted. Secondly, these pre-selected variables were subjected to a forward stepwise variable selection process, adding one variable to the model in every step (i.e., model with 1 variable in step 1, then added one more in each subsequent step). Candidate variables (those not yet selected) were ranked for each step based on the Akaike Information Criterion (AIC) values, where the variable corresponding to the lowest AIC was selected. A second criteria to add the variable to the model was a significant improvement according to a likelihood-ratio test (LRT) with a threshold p-value of 0.05. The variable selection procedure was repeated 5000 times on bootstrapped samples of the training datasets.

The final model performance was evaluated using several metrics. Model fit was assessed by the AIC and Bayesian Information Criterion (BIC). Discriminative ability was quantified using the Receiver Operating Characteristic (ROC) curve with the Area Under the Curve (AUC) and its 95% Confidence Interval (CI). The Nagelkerke R^2^ measured the explained variance, while calibration was evaluated through calibration plots, the Brier score and Log Loss.

### 2.4. Sub-cohort analyses

Radiation dose distributions and treatment protocols are typically tumour site-specific. The distribution of dose across the different subsites varies, and, in addition, the standard primary treatment for locally advanced OCC is surgery followed by post-operative RT (PORT) sometimes combined with concurrent chemotherapy. In contrast, other subsites are predominately treated with primary RT, often combined with concurrent chemotherapy. To account for this, the following sub-cohort analyses were performed for i) a subset of subjects including OPC and locally advanced (T3/T4) larynx/hypopharynx cancer cases, ii) a subset including OCC cases only, iii) a subset including subjects treated with primary RT, and iv) a subset of subjects treated with PORT.

## 3. Results

### 3.1. Data

The initial cohort consisted of 1,263 HNC patients, of which 415 were ORN cases. Table 1 provides a summary of the clinical and demographic data (institution and dataset-specific cohorts are described in additional tables in Supplement B). After the exclusion of missing data, a final dataset of 1,184 subjects (389 ORN cases) was used for analysis. The training set consisted of 948 subjects (309 ORN cases), with a median (IQR) follow-up time of 4.7 (3.2-6.1) and 4.4 (2.7-5.8) years for the ORN and control groups, respectively. The test dataset consisted of 236 subjects (80 ORN cases), with a median (IQR) follow-up time of 5.0 (3.2-6.0) and 4.1 (3.0-5.5) years, respectively. The external validation cohort consisted of 58 subjects (19 ORN cases), with a median follow-up time of 3.7 (2.4-4.7) and 4.8 (1.7-6.1) years, respectively.

**Table 1.**
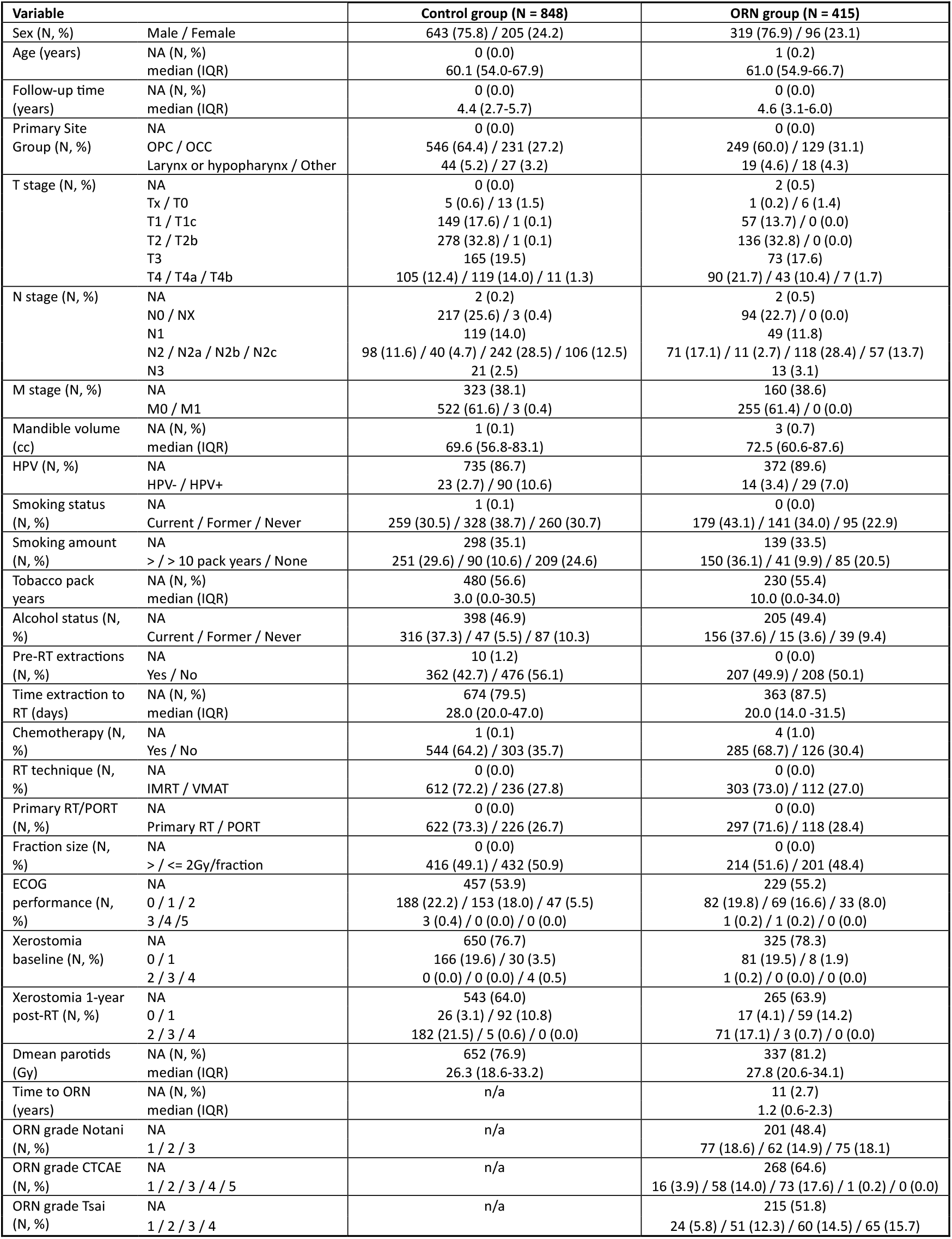
PREDMORN cohort characteristics (see supplementary tables B1 and B2 for subset and institution-specific descriptive tables).

| Variable |  | Control group (N = 848) | ORN group (N = 415) |
| --- | --- | --- | --- |
| Sex (N, %) | Male / Female | 643 (75.8) / 205 (24.2) | 319 (76.9) / 96 (23.1) |
| Age (years) | NA (N, %) | 0 (0.0) | 1 (0.2) |
|  | median (IQR) | 60.1 (54.0-67.9) | 61.0 (54.9-66.7) |
| Follow-up time (years) | NA (N, %) | 0 (0.0) | 0 (0.0) |
|  | median (IQR) | 4.4 (2.7-5.7) | 4.6 (3.1-6.0) |
| Primary Site Group (N, %) | NA | 0 (0.0) | 0 (0.0) |
|  | OPC / OCC | 546 (64.4) / 231 (27.2) | 249 (60.0) / 129 (31.1) |
|  | Larynx or hypopharynx / Other | 44 (5.2) / 27 (3.2) | 19 (4.6) / 18 (4.3) |
| T stage (N, %) | NA | 0 (0.0) | 2 (0.5) |
|  | Tx / T0 | 5 (0.6) / 13 (1.5) | 1 (0.2) / 6 (1.4) |
|  | T1 / T1c | 149 (17.6) / 1 (0.1) | 57 (13.7) / 0 (0.0) |
|  | T2 / T2b | 278 (32.8) / 1 (0.1) | 136 (32.8) / 0 (0.0) |
|  | T3 | 165 (19.5) | 73 (17.6) |
|  | T4 / T4a / T4b | 105 (12.4) / 119 (14.0) / 11 (1.3) | 90 (21.7) / 43 (10.4) / 7 (1.7) |
| N stage (N, %) | NA | 2 (0.2) | 2 (0.5) |
|  | N0 / NX | 217 (25.6) / 3 (0.4) | 94 (22.7) / 0 (0.0) |
|  | N1 | 119 (14.0) | 49 (11.8) |
|  | N2 / N2a / N2b / N2c | 98 (11.6) / 40 (4.7) / 242 (28.5) / 106 (12.5) | 71 (17.1) / 11 (2.7) / 118 (28.4) / 57 (13.7) |
|  | N3 | 21 (2.5) | 13 (3.1) |
| M stage (N, %) | NA | 323 (38.1) | 160 (38.6) |
|  | M0 / M1 | 522 (61.6) / 3 (0.4) | 255 (61.4) / 0 (0.0) |
| Mandible volume (cc) | NA (N, %) | 1 (0.1) | 3 (0.7) |
|  | median (IQR) | 69.6 (56.8-83.1) | 72.5 (60.6-87.6) |
| HPV (N, %) | NA | 735 (86.7) | 372 (89.6) |
|  | HPV- / HPV+ | 23 (2.7) / 90 (10.6) | 14 (3.4) / 29 (7.0) |
| Smoking status (N, %) | NA | 1 (0.1) | 0 (0.0) |
|  | Current / Former / Never | 259 (30.5) / 328 (38.7) / 260 (30.7) | 179 (43.1) / 141 (34.0) / 95 (22.9) |
| Smoking amount (N, %) | NA | 298 (35.1) | 139 (33.5) |
|  | > / > 10 pack years / None | 251 (29.6) / 90 (10.6) / 209 (24.6) | 150 (36.1) / 41 (9.9) / 85 (20.5) |
| Tobacco pack years | NA (N, %) | 480 (56.6) | 230 (55.4) |
|  | median (IQR) | 3.0 (0.0-30.5) | 10.0 (0.0-34.0) |
| Alcohol status (N, %) | NA | 398 (46.9) | 205 (49.4) |
|  | Current / Former / Never | 316 (37.3) / 47 (5.5) / 87 (10.3) | 156 (37.6) / 15 (3.6) / 39 (9.4) |
| Pre-RT extractions (N, %) | NA | 10 (1.2) | 0 (0.0) |
|  | Yes / No | 362 (42.7) / 476 (56.1) | 207 (49.9) / 208 (50.1) |
| Time extraction to RT (days) | NA (N, %) | 674 (79.5) | 363 (87.5) |
|  | median (IQR) | 28.0 (20.0-47.0) | 20.0 (14.0 -31.5) |
| Chemotherapy (N, %) | NA | 1 (0.1) | 4 (1.0) |
|  | Yes / No | 544 (64.2) / 303 (35.7) | 285 (68.7) / 126 (30.4) |
| RT technique (N, %) | NA | 0 (0.0) | 0 (0.0) |
|  | IMRT / VMAT | 612 (72.2) / 236 (27.8) | 303 (73.0) / 112 (27.0) |
| Primary RT/PORT (N, %) | NA | 0 (0.0) | 0 (0.0) |
|  | Primary RT / PORT | 622 (73.3) / 226 (26.7) | 297 (71.6) / 118 (28.4) |
| Fraction size (N, %) | NA | 0 (0.0) | 0 (0.0) |
|  | > / <= 2Gy/fraction | 416 (49.1) / 432 (50.9) | 214 (51.6) / 201 (48.4) |
| ECOG performance (N, %) | NA | 457 (53.9) | 229 (55.2) |
|  | 0 / 1 / 2 | 188 (22.2) / 153 (18.0) / 47 (5.5) | 82 (19.8) / 69 (16.6) / 33 (8.0) |
|  | 3 / 4 / 5 | 3 (0.4) / 0 (0.0) / 0 (0.0) | 1 (0.2) / 1 (0.2) / 0 (0.0) |
| Xerostomia baseline (N, %) | NA | 650 (76.7) | 325 (78.3) |
|  | 0 / 1 | 166 (19.6) / 30 (3.5) | 81 (19.5) / 8 (1.9) |
|  | 2 / 3 / 4 | 0 (0.0) / 0 (0.0) / 4 (0.5) | 1 (0.2) / 0 (0.0) / 0 (0.0) |
| Xerostomia 1-year post-RT (N, %) | NA | 543 (64.0) | 265 (63.9) |
|  | 0 / 1 | 26 (3.1) / 92 (10.8) | 17 (4.1) / 59 (14.2) |
|  | 2 / 3 / 4 | 182 (21.5) / 5 (0.6) / 0 (0.0) | 71 (17.1) / 3 (0.7) / 0 (0.0) |
| Dmean parotids (Gy) | NA (N, %) | 652 (76.9) | 337 (81.2) |
|  | median (IQR) | 26.3 (18.6-33.2) | 27.8 (20.6-34.1) |
| Time to ORN (years) | NA (N, %) | n/a | 11 (2.7) |
|  | median (IQR) |  | 1.2 (0.6-2.3) |
| ORN grade Notani (N, %) | NA | n/a | 201 (48.4) |
|  | 1 / 2 / 3 |  | 77 (18.6) / 62 (14.9) / 75 (18.1) |
| ORN grade CTCAE (N, %) | NA | n/a | 268 (64.6) |
|  | 1 / 2 / 3 / 4 / 5 |  | 16 (3.9) / 58 (14.0) / 73 (17.6) / 1 (0.2) / 0 (0.0) |
| ORN grade Tsai (N, %) | NA | n/a | 215 (51.8) |
|  | 1 / 2 / 3 / 4 |  | 24 (5.8) / 51 (12.3) / 60 (14.5) / 65 (15.7) |

#### Clinical data

The dominating primary tumour site was OPC (63.0%), followed by OCC (28.5%), larynx/hypopharynx (5.0%) and other sites (3.6%). While tobacco smoking status was included as a variable for statistical analyses with observed statistical association to ORN (p=0.002) at univariable analysis (see supplementary Table C1), the amount of tobacco (tobacco pack years) was not uniformly well recorded. However, based on a subset of 368 controls and 185 ORN subjects, the median (IQR) of pack years for ORN exceeded that of the controls: 10.0 (0.0-34.0) vs. 3.0 (0.0-30.5), although this difference was not statistically significant (p=0.094). Dental extractions pre-RT were performed in a higher percentage within the ORN group (49.9% vs. 42.7%) with a statistically significant association with ORN at univariable analysis (p=0.026). Although information on the elapsed time between dental extractions and the start of RT was largely missing, on a subset of 174 controls and 52 ORN cases with available data, the median (IQR) time was shorter for ORN cases: 20.0 days (14.0-31.5) vs. 28.0 days (20.0-47.0) and this difference was found statistically significant (p=0.002). Chemotherapy was more common in ORN cases (68.7% vs. 64.2%) and was statistically associated with this endpoint (p= 0.042). Xerostomia at one-year post-RT was recorded for 455 (36.0%) subjects (305 controls and 150 ORN), and a significantly (p = 0.015) higher percentage of grade ≥ 2 xerostomia was observed for the control group (22.1% vs. 17.8%). Mandible volume varied largely across institutions (supplementary Figure D1) and was significantly associated with ORN at univariable analysis (p=0.013) with ORN cases exhibiting larger volumes, 72.5 cc (60.6-87.6) vs. 69.6 (56.8-83.1). Mandible volume, however, was significantly associated to PORT status (t-test p-value < 0.001), with smaller mandible volumes observed in patients who underwent surgery prior RT (i.e., PORT positive status) (median 64.4 cc, IQR 52.3-75.3) compared to patients receiving RT as their primary treatment (73.1 cc, 60.6-87.0) (see supplementary Figure D2).

#### Dosimetric data

The median cumulative mandibular DVH was higher for the ORN group than for the controls as shown in Figure 1; this was also true for individual institution-specific DVH data (see supplementary Figure E1). In particular, the median (IQR) V_50Gy_ to the mandible was 45.8% (31.8-61.6) and 34.1% (23.3-50.2) for the ORN and control groups, respectively (p<0.0001); the median D_2%_ to the mandible was 67.6 Gy (63.4-71.3) and 66.3 Gy (62.2-69.4) for the ORN and control groups, respectively (p<0.0001). On univariable analysis for the entire PREDMORN cohort, most DVH metrics were significantly related to ORN (p<0.05), except for V_5Gy,_ V_10Gy_, V_15Gy_, D_90%_, D_95%_ and D_98%_; a smaller set of significant DVH metrics was observed on the OCC sub-cohort (see supplementary Table C1). Parotid gland doses were available in 21.7% of subjects (196 controls and 78 ORN cases), with mean doses for the ORN group marginally higher than for the control group, with a median (IQR) of 27.8 Gy (20.6-34.1) vs. 26.3 (18.6-33.2).

**Figure 1.**
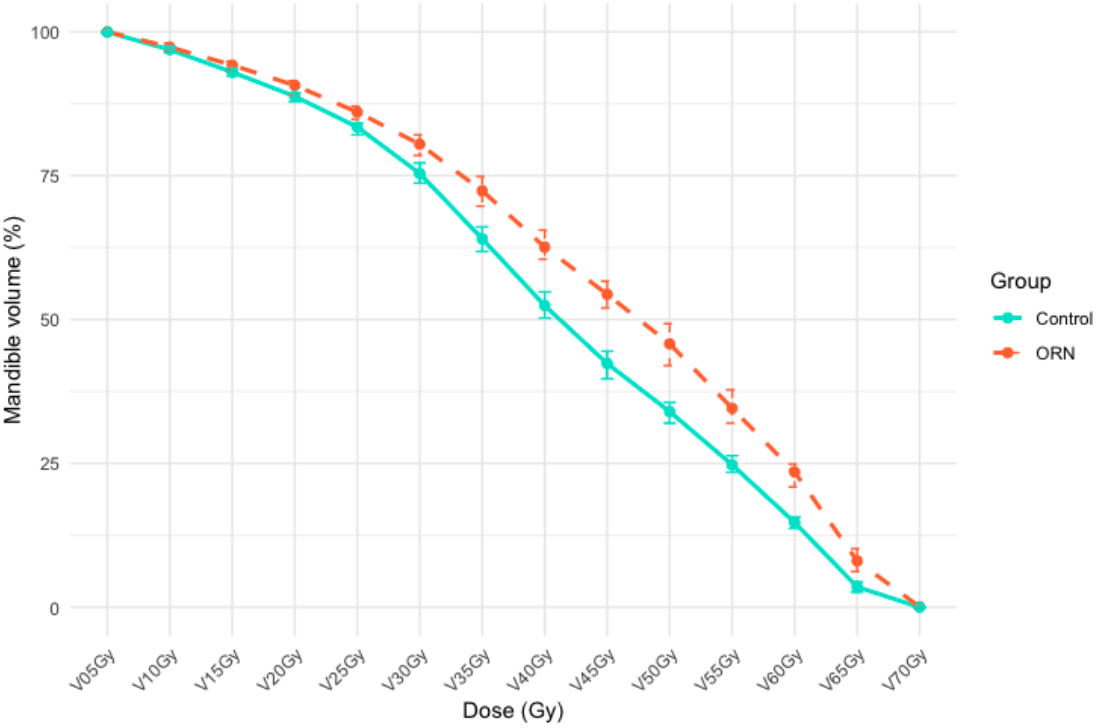
Median cumulative DVH plots for the ORN and control groups of the PREDMORN cohort. The medians were calculated for each DVH metric across the ORN and control groups. Error bars correspond to the 95% confidence intervals of the medians estimated using the binomial method, which identifies the lower and upper ranks corresponding to the 95% confidence bounds based on the size and distribution of the data. Additional institution-specific DVH plots and DVH plots for the OPC/advanced larynx cancer and OCC sub-cohorts are provided in Supplement E.

**Figure 2.**
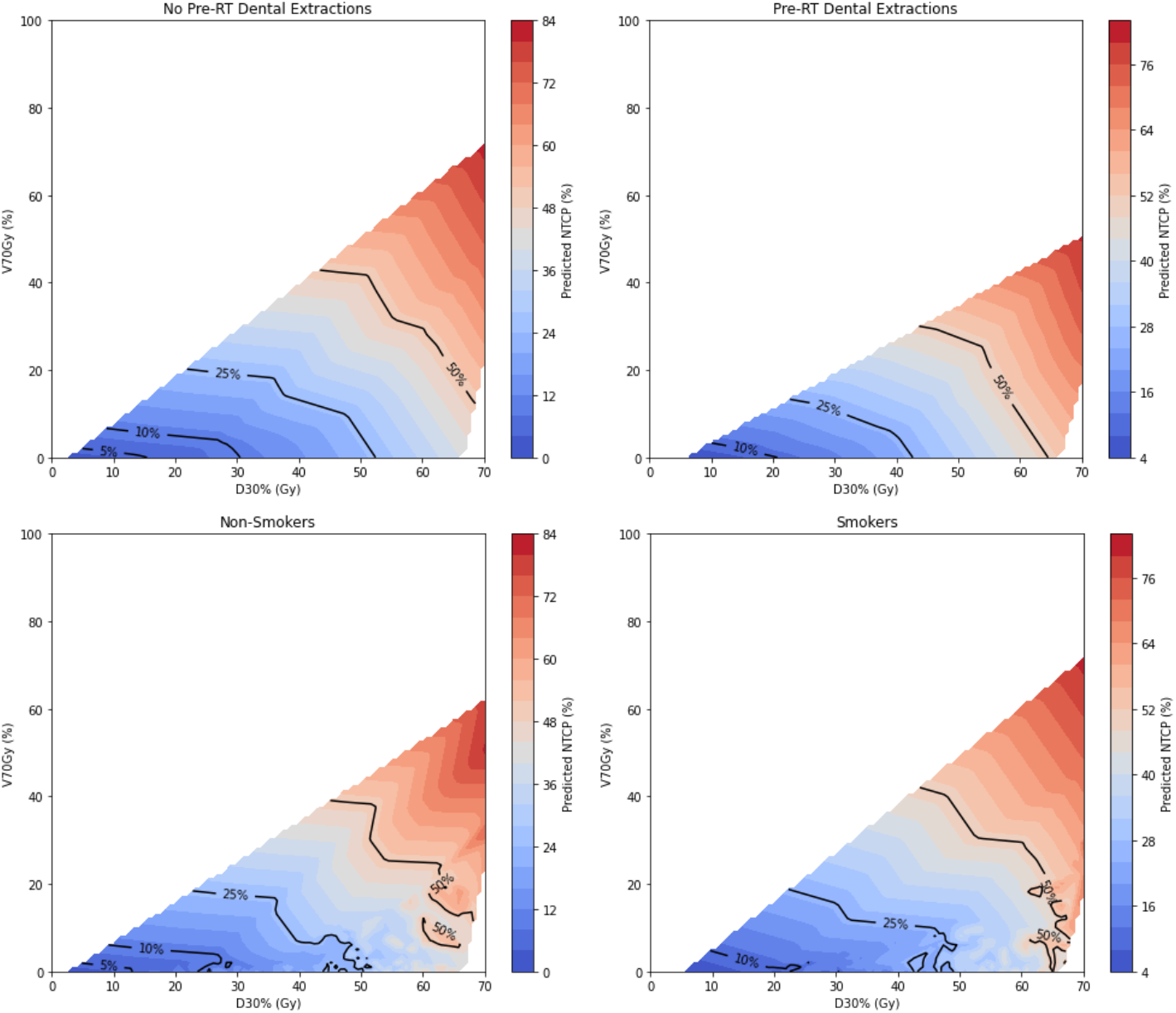
NTCP (Normal-Tissue Complication Probability) contour plots for mandibular osteoradionecrosis (ORN) risk as a function of D30% and V70Gy, stratified by a) pre-RT dental extractions and b) smoking status. The NTCP values were interpolated across the dose range for visualization.

#### Clinical endpoint

The median (IQR) time from the end of RT to ORN diagnosis was 1.2 years (0.6-2.3 with a range across centres of 0.6 to 7.4 years (see supplementary Figure F1). A statistically significant difference in time to ORN curves was observed between patients with and without pre-RT dental extractions (p = 0.002) and between patients with and without a history of smoking (p = 0.012). Patients who underwent pre-RT dental extractions or with a positive smoking status (i.e., current smoker) showed a higher probability of developing ORN earlier, with an increase of up to 18.7% and 13.2% at 0.8 and 3.2 years, respectively (see supplementary figures F2a and F2b, respectively). Of all 415 ORN cases, 140 (33.7%) were advanced grade ORN (either Notani grade III or Tsai grade 4).

### 3.2. Variable pre-selection

All 8 clinical variables considered for multivariable statistical analysis – Pre-RT dental extractions, smoking status, chemotherapy, fraction size, sex, RT technique, PORT and age – were pre-selected based on their low collinearity, as determined by the Spearman correlation test (supplementary Figure G1 and Table G1). This outcome was anticipated, given that the proposed set of clinical variables was intentionally designed to capture potential clinical risk factors without redundancy. Dose-volume variables, however, are inherently correlated because they originate from a single source (DVH). Consequently, during the multicollinearity reduction process, only a subset of 5 dose-volume variables (D_10%_, D_30%_, D_60%_, V_25Gy_ and V_70Gy_) was selected for further analysis.

### 3.3. Multivariable ORN NTCP_generic_ model

The forward stepwise selection process (supplementary Table G2) identified the following variables as the best set of predictors for ORN in the following order of importance: D_30%_, pre-RT dental extraction, V_70Gy_ and smoking status. A low correlation was observed between the two dosimetric factors selected, D_30%_ and V_70Gy_ (supplementary Figure G2). These variables remained the most frequently selected in the bootstrapped analyses (supplementary Figure G3). Positive regression coefficients were obtained for all model parameters (Table 2), indicating that an increase in these risk factors resulted in a higher mandibular ORN risk. Refer to Supplement I for a detailed explanation on the practical application of this NTCP model.

**Table 2:**
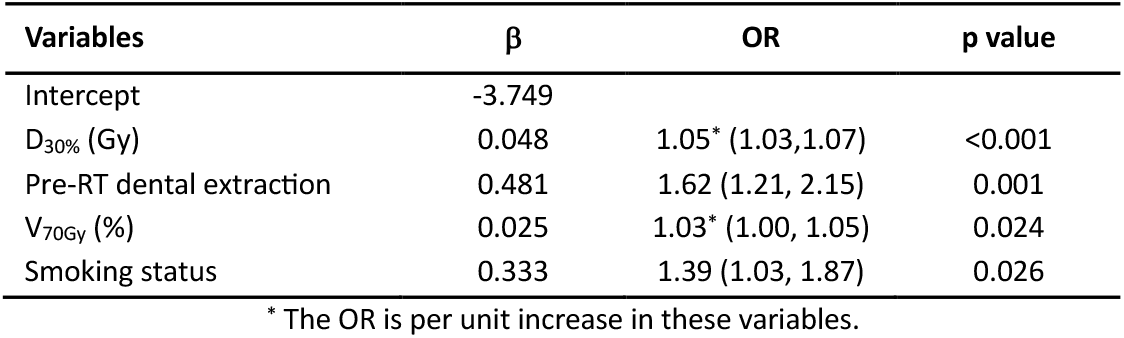
ORN NTCP model parameters (see Supplement I for guidance on the practical use of this model).

| Variables | $\beta$ | OR | p value |
| --- | --- | --- | --- |
| Intercept | -3.749 |  |  |
| D<sub>30%</sub> (Gy) | 0.048 | 1.05* (1.03,1.07) | <0.001 |
| Pre-RT dental extraction | 0.481 | 1.62 (1.21, 2.15) | 0.001 |
| V<sub>70Gy</sub> (%) | 0.025 | 1.03* (1.00, 1.05) | 0.024 |
| Smoking status | 0.333 | 1.39 (1.03, 1.87) | 0.026 |

\* The OR is per unit increase in these variables.

### 3.4. ORN NTCP model performance

The ORN NTCP model demonstrated moderate overall performance with AUCs of 0.67 (95% CI: 0.64-0.71), 0.69 (0.62-0.76) and 0.69 (0.53-0.84) on the train, test and external validation datasets, respectively (Table 3). Calibration plots demonstrate good model calibration in both the train (Figure 3a) and the two validation cohorts (figures 3b and 3c), as the predicted and observed rates matched the identity line closely (intercept range −0.03-0.00; slope range: 0.99-1.09).

**Table 3.** Model performance results of the ORN NTCP model on the PREDMORN cohort. The 95% confidence interval (CI) for the area under the ROC curve (AUC) has been calculated using the DeLong method.

| ORN NTCP model performance |  |  |  |
| --- | --- | --- | --- |
|  | Training (N=948) | Test (N=236) | External validation (N=58) |
| ROC AUC (95% CI) | 0.67 (0.64-0.71) | 0.69 (0.62-0.76) | 0.69 (0.53-0.84) |
| Nagelkerke R<sup>2</sup> | 0.108 | 0.122 | 0.098 |
| Brier score | 0.203 | 0.204 | 0.199 |
| Log Loss | 0.591 | 0.594 | 0.596 |

**Figure 3.**
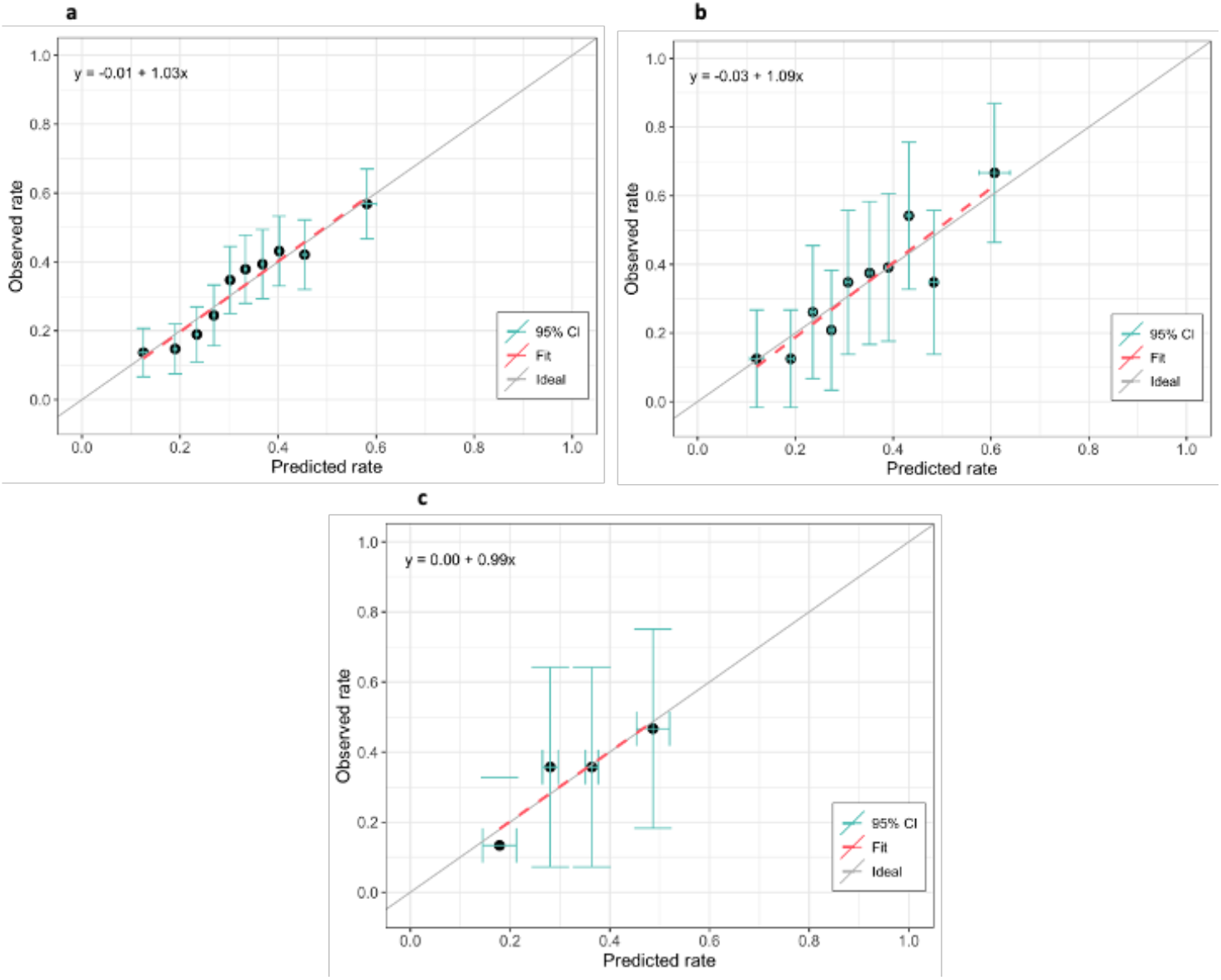
Calibration curves when testing the NTCP model on the a) train, b) test, and c) external validation datasets. The number of bins in the calibration plots was adjusted in each plot based on dataset size for each cohort/sub-cohort to optimise representation of the data (i.e., 10 bins for the train and test plots, 4 bins for the external validation plot).

### 3.5. Sub-cohort analyses

When applied to the OPC/locally advanced (T3/T4) larynx/hypopharynx cancer sub-cohort, the model’s performance improved (see supplemental Table H1), with ROC AUC values increasing to 0.75 (for both the test and external validation datasets), and higher Nagelkerke R^2^ values, indicating better discrimination and fit. The Brier scores and Log Loss also improved, reflecting enhanced accuracy in this sub-cohort. Similarly, the model’s performance was higher when tested on the Primary RT sub-cohort, with ROC AUC values of 0.67 and 0.72 for testing and external validation, respectively (see supplementary Table H2). Conversely, the model’s performance decreased when tested on the OCC or PORT sub-cohorts.

## 4. Discussion

In this first analysis performed by the PREDMORN Consortium, we have aimed to reproduce existing statistical association and modelling analyses on the largest and most diverse multi-institutional mandibular ORN cohort worldwide to allow comparison with previous studies. As such, we have developed, tested and externally validated an NTCP model for ORN.

On a cohort of 1,184 subjects with 389 ORN cases, the multivariable ORN NTCP model identified key risk factors as D_30%_ (the dose received by 30% of the mandible volume), pre-RT dental extractions, V_70Gy_ (the mandible volume receiving at least 70 Gy) and smoking status. Our model showed a moderate predictive performance (AUC 0.69 both for testing and external validation), but improved accuracy in OPC and locally advanced (T3/T4) larynx/hypopharynx (AUC 0.75 for both testing and external validation) and primary RT (AUC 0.70 and 0.72) sub-cohorts.

The dosimetric variable D_30%_ was selected by the NTCP model. The median D_30%_ (IQR) across the ORN and control groups was 57.4 Gy (51.4-61.9) and 52.5 Gy (45.1-71.9), respectively. In van Dijk et al.^7^, D_30%_ was also selected in their NTCP models, with thresholds of 35 Gy and 42 Gy for a <5% risk of ORN in patients with and without pre-dental extractions, respectively. Our model identified slightly lower thresholds, with doses below 20 Gy for non-smokers or no extractions and below 10 Gy otherwise. However, our model was developed on a 2:1 match cohort, with an artificially elevated ORN incidence rate of 32.9%, and thus the reported ORN risk probabilities cannot be directly compared to those from the population cohort used in van Dijk et al.^7^, with an incidence ORN rate of 13.7%. A such, the probabilities predicted by our NTCP model should be considered as relative rather than absolute^17^ and would need to be adjusted in order to obtain real ORN risk probabilities based on the baseline ORN risk of the target population^18^ (see Supplement I for a worked example).

Higher radiation doses are a known risk factor for ORN and the 2024 ISOO-MASCC-ASCO guidelines^19^ state that patients receiving radiation doses to the mandible higher than 50 Gy should be considered at risk of developing ORN. Previous studies, however, have also reported differences between the ORN and control groups at low doses^10^ and suggested microvasculature collapse in the mandible at doses around 30 Gy^20^. Our international pooled data analysis suggests that even lower doses should be considered in treatment planning to minimise the risk of ORN, with thresholds as described in the preceding paragraph, in addition to supporting a D_30%_ of 35-40 Gy^7^ or a D_mean_ of 30 Gy^21^ as previously reported.

The risk of ORN escalates with dental extractions or surgery, as the compromised vascularisation in an irradiated mandible impairs the mandible’s healing capacity after such procedures, where the time between dental extraction and the start of RT to allow for the healing process plays a crucial role^22^. The ISOO-MASCC-ASCO guidelines^19^ recommend a ‘2-week healing period between the time of dental extraction and the start of radiation therapy’; most participating PREDMORN institutions met this recommendation, with slightly overall shorter times observed in ORN cases (20.0 vs. 28.0 days). In line with previous work^9^, dental extraction pre-RT was observed to negatively impact the longitudinal risk of developing ORN. Further investigation should focus on a more complete dataset of time from extractions to RT start and the association between extraction site and ORN localisation.

As highlighted in previous studies^20,23,24^, there is an increased vulnerability to ORN in the HPV-associated OPC group of patients, who are generally younger, with better dental status and more often without the lifestyle factors traditionally associated with ORN such as smoking^25^, challenging the stigma that HNC patients have neglected and poor dentition^26^. However, we found similar smoking rates (p-value=0.91) in the OPC subgroup (28.3% and 42.4% for the control and ORN groups, respectively) compared to other primary sites (30.5% and 44.6% for the control and ORN groups).

Patients with HNC, mostly OCC cases, may undergo pre-RT mandibular surgery, such as rim resection or hemimandibulectomy. For the PREDMORN cohort, 81.2% of OCC cases were treated with PORT, where the whole mandible is typically irradiated to a more homogeneous dose compared to OPC or locally advanced larynx/hypopharynx cases. This results in lower discriminative potential for the DVH variables, especially in lower and intermediate doses (as shown in supplementary Figure E2b), which could have contributed to a worse model performance for the OCC and PORT sub-cohorts.

A recent study by Verduijn et al.^27^ also observed an association between mandibular volume and ORN and speculated that this could be related to the dentition status of the patient. In our study, mandible volume was found to be statistically associated with ORN at univariable analysis for the OCC sub-cohort (p-value 0.024) but not for the OPC/locally advanced larynx/hypopharynx sub-cohort (p-value 0.079). On the other hand, mandible volume was found to be statistically associated with PORT status, with smaller mandible volumes observed in patients treated with PORT (median 64.4 cc, IQR 52.3-75.3) vs. primary RT (73.1 cc, 60.6-87.0) (see supplementary Figure D2). Further investigation is required concerning this less explored risk factor. The irradiation of a partially resected mandible may contribute to the risk of ORN due to the combined effects of surgery and radiation compromising its vascularisation. Building on previous studies^5,28,29^ that have characterised imaging parameters linked to vascular damage in the irradiated mandible, future research should leverage imaging data to further elucidate the interplay between pre-RT surgery, mandibular volume, and ORN incidence.

This study has some limitations. A matched control-case approach based on primary tumour site was followed as whole population consecutive datasets were not available from all participating institutions; hence, we do not know the incidence of ORN across the populations from which the cases and controls were extracted. This potentially introduced bias in the representation of the tumour sites within the cohort both across participating institutions and when compared with previous studies (e.g., the proportion of larynx cases we considered was smaller than in the population-based cohort considered in van Dijk et al.^7^). Differences in case mix and ORN incidence rate (i.e., natural ORN incidence in a consecutive dataset compared to that in a 2:1 matched cohort) could account for the superiority of their model’s discrimination performance.

The choice of dose reporting formalism—whether D_w,w_, D_w,m_ or D_m,m_ —varied across institutions, where D_x,y_ represents the dose to medium x (w for water or m for other medium) calculated using the energy deposition in medium y. This variability likely introduced a degree of systematic uncertainty in the DVH values, particularly in high-density regions such as bone or metal implants^30^. As this study was conducted retrospectively, dose recalculation was not feasible; however, we recommend establishing a standardised dose reporting framework for future studies to ensure consistency and comparability across institutions.

Acknowledging the different stages or grades of ORN is highly relevant as the clinical intervention required will often depend on the severity of ORN, with the more severe ORN involving bone fracture requiring surgical intervention. Because of the lack of consensus in the classification system for ORN, in this study, we dichotomised the endpoint (ORN vs. no ORN) rather than being able to consider separate stages. A new staging system, ClinRad, has been recently proposed by Watson et al.^6^ that reportedly outperforms existing systems and has been endorsed for prospective use by ISOO-MASCC-ASCO guidelines^19^ to standardize reporting on ORN.

Existing efforts in modelling ORN have used limited size and single institution datasets^7^ and lacked recommended external validation^31^, potentially leading to overoptimistic single-institution models. The low prevalence of ORN represents a statistical challenge that limits trust and the successful adoption of existing research findings in the clinic. The existence of multiple coexisting ORN classification systems adds complexity to multi-institutional efforts^32^. While the PREDMORN consortium was developed to address these limitations, and the present study represents a significant advancement in this direction, we acknowledge that further validation of our results is required. Specifically, the size of the dataset used for external validation was small the reliability of our results. While data handling alternatives (e.g., mixing and shuffling datasets) could have potentially improved our results, we tried to maintain research rigour by following the planned protocol^14^. Instead, we will aim to repeat the external validation process in additional multiple independent cohorts.

Finally, the present work adopted a more traditional approach to modelling ORN to establish a baseline that enables comparison to existing publications. The PREDMORN study has been planned in two phases: while the first phase has used DVH data, the second phase will focus on higher dimensionality (imaging) data to incorporate spatial information. Future work will also explore other statistical options that do not assume linearity and address the inherent multicollinearity of DVH-based parameters^33^.

## Data Availability

All data produced in this multi-centre study are available upon reasonable request and subject to a Material Transfer Agreement (MTA) with each participating institution individually.

## References

(1) Frankart, A. J.; Frankart, M. J.; Cervenka, B.; Tang, A. L.; Krishnan, D. G.; Takiar, V. Osteoradionecrosis: Exposing the Evidence Not the Bone. Int. J. Radiat. Oncol. Biol. Phys. 2021, 109 (5), 1206–1218. 10.1016/j.ijrobp.2020.12.043.

(2) van der Laan, H. P.; Van den Bosch, L.; Schuit, E.; Steenbakkers, R. J. H. M.; van der Schaaf, A.; Langendijk, J. A. Impact of Radiation-Induced Toxicities on Quality of Life of Patients Treated for Head and Neck Cancer. Radiother. Oncol. 2021, 160, 47–53. 10.1016/j.radonc.2021.04.011.

(3) Patel, V.; Ormondroyd, L.; Lyons, A.; McGurk, M. The Financial Burden for the Surgical Management of Osteoradionecrosis. Br. Dent. J. 2017, 222 (3), 177–180. 10.1038/sj.bdj.2017.121.

(4) Marx, R. E. Osteoradionecrosis: A New Concept of Its Pathophysiology. J. Oral Maxillofac. Surg. 1983, 41 (5), 283–288. 10.1016/0278-2391(83)90294-X.

(5) Mohamed, A. S. R.; He, R.; Ding, Y.; Wang, J.; Fahim, J.; Elgohari, B.; Elhalawani, H.; Kim, A. D.; Ahmed, H.; Garcia, J. A.; Johnson, J. M.; Stafford, R. J.; Bankson, J. A.; Chambers, M. S.; Sandulache, V. C.; Fuller, C. D.; Lai, S. Y. Quantitative Dynamic Contrast-Enhanced MRI Identifies Radiation-Induced Vascular Damage in Patients With Advanced Osteoradionecrosis: Results of a Prospective Study. Int. J. Radiat. Oncol. Biol. Phys. 2020, 108 (5), 1319–1328. 10.1016/j.ijrobp.2020.07.029.

(6) Watson, E. E.; Hueniken, K.; Lee, J.; Huang, S. H.; Maghrabi A A. E.,; Xu, W.; Moreno, A. C.; Tsai, C. J.; Hahn, E.; McPartlin, A. J.; Yao, C. M.; Goldstein, D. P.; De Almeida, J. R.; Waldon, J. N.; Fuller, C. D.; Hope, A. J.; Ruggiero, S. L.; Glogauer, M.; Hosni, A. A. Development and Standardization of a Classification System for Osteoradionecrosis: Implementation of a Risk-Based Model. MedRxiv Prepr. Serv. Health Sci. 2023, 2023.09.12.23295454. 10.1101/2023.09.12.23295454.

(7) van Dijk, L. V.; Abusaif, A. A.; Rigert, J.; Naser, M. A.; Hutcheson, K. A.; Lai, S. Y.; Fuller, C. D.; Mohamed, A. S. R. Normal Tissue Complication Probability (NTCP) Prediction Model for Osteoradionecrosis of the Mandible in Patients With Head and Neck Cancer After Radiation Therapy: Large-Scale Observational Cohort. Int. J. Radiat. Oncol. 2021, 111 (2), 549–558. 10.1016/j.ijrobp.2021.04.042.

(8) Habib, S.; Sassoon, I.; Thompson, I.; Patel, V. Risk Factors Associated with Osteoradionecrosis. Oral Surg. 2021, 14 (3), 227–235. 10.1111/ors.12597.

(9) Group, M. A. H. and N. S. W.; Humbert-Vidan, L.; Kamel, S.; Wentzel, A.; Kaffey, Z.; Abdelaal, M.; Spier, K. B.; West, N. A.; Marai, G. E.; Canahuate, G.; Zhang, X.; Chen, M. M.; Wahid, K. A.; Rigert, J.; Hosseinian, S.; Schaefer, A. J.; Brock, K. K.; Chambers, M.; Otun, A. O.; Aponte-Wesson, R.; Patel, V.; Hope, A.; Phan, J.; Garden, A. S.; Frank, S. J.; Morrison, W. H.; Spiotto, M. T.; Rosenthal, D.; Lee, A.; He, R.; Naser, M. A.; Watson, E.; Hutcheson, K. A.; Mohamed, A. S. R.; Sandulache, V. C.; Dijk, L. V. van; Moreno, A. C.; Urbano, T. G.; Lai, S. Y.; Fuller, C. D. Mandibular Dose-Volume Predicts Time-to-Osteoradionecrosis in an Actuarial Normal-Tissue Complication Probability (NTCP) Model: External Validation of Right-Censored Clinico-Dosimetric and Competing Risk Application across International Multi-Institutional Observational Cohorts and Online Graphical User Interface Clinical Support Tool Assessment. medRxiv August 20, 2024, p 2024.08.20.24312311.

(10) Aarup-Kristensen, S.; Hansen, C. R.; Forner, L.; Brink, C.; Eriksen, J. G.; Johansen, J. Osteoradionecrosis of the Mandible after Radiotherapy for Head and Neck Cancer: Risk Factors and Dose-Volume Correlations. Acta Oncol. 2019, 58 (10), 1373–1377. 10.1080/0284186X.2019.1643037.

(11) Möring, M. M.; Mast, H.; Wolvius, E. B.; Verduijn, G. M.; Petit, S. F.; Sijtsema, N. D.; Jonker, B. P.; Nout, R. A.; Heemsbergen, W. D. Osteoradionecrosis after Postoperative Radiotherapy for Oral Cavity Cancer: A Retrospective Cohort Study. Oral Oncol. 2022, 133, 106056. 10.1016/j.oraloncology.2022.106056.

(12) Kubota, H.; Miyawaki, D.; Mukumoto, N.; Ishihara, T.; Matsumura, M.; Hasegawa, T.; Akashi, M.; Kiyota, N.; Shinomiya, H.; Teshima, M.; Nibu, K.; Sasaki, R. Risk Factors for Osteoradionecrosis of the Jaw in Patients with Head and Neck Squamous Cell Carcinoma. Radiat. Oncol. 2021, 16 (1), 1. 10.1186/s13014-020-01701-5.

(13) Lang, K.; Held, T.; Meixner, E.; Tonndorf-Martini, E.; Ristow, O.; Moratin, J.; Bougatf, N.; Freudlsperger, C.; Debus, J.; Adeberg, S. Frequency of Osteoradionecrosis of the Lower Jaw after Radiotherapy of Oral Cancer Patients Correlated with Dosimetric Parameters and Other Risk Factors. Head Face Med. 2022, 18, 7. 10.1186/s13005-022-00311-8.

(14) Humbert-Vidan, L.; Hansen, C. R.; Fuller, C. D.; Petit, S.; van der Schaaf, A.; van Dijk, L. V.; Verduijn, G. M.; Langendijk, H.; Muñoz-Montplet, C.; Heemsbergen, W.; Witjes, M.; Mohamed, A. S. R.; Khan, A. A.; Marruecos Querol, J.; Oliveras Cancio, I.; Patel, V.; King, A. P.; Johansen, J.; Guerrero Urbano, T. Protocol Letter: A Multi-Institutional Retrospective Case-Control Cohort Investigating PREDiction Models for Mandibular OsteoRadioNecrosis in Head and Neck Cancer (PREDMORN). Radiother. Oncol. 2022, 176, 99–100. 10.1016/j.radonc.2022.09.014.

(15) Notani, K.; Yamazaki, Y.; Kitada, H.; Sakakibara, N.; Fukuda, H.; Omori, K.; Nakamura, M. Management of Mandibular Osteoradionecrosis Corresponding to the Severity of Osteoradionecrosis and the Method of Radiotherapy. Head Neck 2003, 25 (3), 181–186. 10.1002/hed.10171.

(16) Tsai, C. J.; Hofstede, T. M.; Sturgis, E. M.; Garden, A. S.; Lindberg, M. E.; Wei, Q.; Tucker, S. L.; Dong, L. Osteoradionecrosis and Radiation Dose to the Mandible in Patients with Oropharyngeal Cancer. Int. J. Radiat. Oncol. Biol. Phys. 2013, 85 (2), 415–420. 10.1016/j.ijrobp.2012.05.032.

(17) Moons, K. G. M.; Kengne, A. P.; Woodward, M.; Royston, P.; Vergouwe, Y.; Altman, D. G.; Grobbee, D. E. Risk Prediction Models: I. Development, Internal Validation, and Assessing the Incremental Value of a New (Bio)Marker. Heart 2012, 98 (9), 683–690. 10.1136/heartjnl-2011-301246.

(18) Moons, K. G. M.; Kengne, A. P.; Grobbee, D. E.; Royston, P.; Vergouwe, Y.; Altman, D. G.; Woodward, M. Risk Prediction Models: II. External Validation, Model Updating, and Impact Assessment. Heart Br. Card. Soc. 2012, 98 (9), 691–698. 10.1136/heartjnl-2011-301247.

(19) Peterson, D. E.; Koyfman, S. A.; Yarom, N.; Lynggaard, C. D.; Ismaila, N.; Forner, L. E.; Fuller, C. D.; Mowery, Y. M.; Murphy, B. A.; Watson, E.; Yang, D. H.; Alajbeg, I.; Bossi, P.; Fritz, M.; Futran, N. D.; Gelblum, D. Y.; King, E.; Ruggiero, S.; Smith, D. K.; Villa, A.; Wu, J. S.; Saunders, D. Prevention and Management of Osteoradionecrosis in Patients With Head and Neck Cancer Treated With Radiation Therapy: ISOO-MASCC-ASCO Guideline. J. Clin. Oncol. Off. J. Am. Soc. Clin. Oncol. 2024, 42 (16), 1975–1996. 10.1200/JCO.23.02750.

(20) Patel, V.; Humbert-Vidan, L.; Thomas, C.; Sassoon, I.; McGurk, M.; Fenlon, M.; Guerrero Urbano, T. Radiotherapy Quadrant Doses in Oropharyngeal Cancer Treated with Intensity Modulated Radiotherapy. Fac. Dent. J. 2020, 11 (4), 166–172. 10.1308/rcsfdj.2020.113.

(21) Aarup-Kristensen, S.; Hansen, C. R.; Forner, L.; Brink, C.; Eriksen, J. G.; Johansen, J. Osteoradionecrosis of the Mandible after Radiotherapy for Head and Neck Cancer: Risk Factors and Dose-Volume Correlations. Acta Oncol. 2019, 58 (10), 1373–1377. 10.1080/0284186X.2019.1643037.

(22) Lee, J.; Hueniken, K.; Cuddy, K.; Pu, J.; El Maghrabi, A.; Hope, A.; Hosni, A.; Glogauer, M.; Watson, E. Dental Extractions Before Radiation Therapy and the Risk of Osteoradionecrosis in Patients With Head and Neck Cancer. JAMA Otolaryngol.--Head Neck Surg. 2023, 149 (12), 1130–1139. 10.1001/jamaoto.2023.3429.

(23) Raguse, J. D.; Hossamo, J.; Tinhofer, I.; Hoffmeister, B.; Budach, V.; Jamil, B.; Jöhrens, K.; Thieme, N.; Doll, C.; Nahles, S.; Hartwig, S. T.; Stromberger, C. Patient and Treatment-Related Risk Factors for Osteoradionecrosis of the Jaw in Patients with Head and Neck Cancer. Oral Surg. Oral Med. Oral Pathol. Oral Radiol. 2016, 121 (3), 215–221.e1. 10.1016/j.oooo.2015.10.006.

(24) Patel, D.; Haria, S.; Patel, V. Oropharyngeal Cancer and Osteoradionecrosis in a Novel Radiation Era: A Single Institution Analysis. Oral Surg. 2021, 14 (2), 113–121. 10.1111/ors.12546.

(25) Lechner, M.; Liu, J.; Masterson, L.; Fenton, T. R. HPV-Associated Oropharyngeal Cancer: Epidemiology, Molecular Biology and Clinical Management. Nat. Rev. Clin. Oncol. 2022, 19 (5), 306. 10.1038/s41571-022-00603-7.

(26) Patel, V.; Kwok, J.; Burke, M.; Urbano, T. G.; Fenlon, M. Should the HPV Positive Oropharyngeal Cancer Patient Be Considered for a Two-Stage Dental Assessment for Their Radiation Treatment? Radiother. Oncol. 2021, 164, 232–235. 10.1016/j.radonc.2021.09.021.

(27) Verduijn, G. M.; Sijtsema, N. D.; van Norden, Y.; Heemsbergen, W. D.; Mast, H.; Sewnaik, A.; Chin, D.; Baker, S.; Capala, M. E.; van der Lugt, A.; van Meerten, E.; Hoogeman, M. S.; Petit, S. F. Accounting for Fractionation and Heterogeneous Dose Distributions in the Modelling of Osteoradionecrosis in Oropharyngeal Carcinoma Treatment. Radiother. Oncol. J. Eur. Soc. Ther. Radiol. Oncol. 2023, 188, 109889. 10.1016/j.radonc.2023.109889.

(28) Joint Head and Neck Radiotherapy-MRI Development Cooperative. Dynamic Contrast-Enhanced MRI Detects Acute Radiotherapy-Induced Alterations in Mandibular Microvasculature: Prospective Assessment of Imaging Biomarkers of Normal Tissue Injury. Sci. Rep. 2016, 6, 29864. 10.1038/srep29864.

(29) Barua, S.; Elhalawani, H.; Volpe, S.; Al Feghali, K. A.; Yang, P.; Ng, S. P.; Elgohari, B.; Granberry, R. C.; Mackin, D. S.; Gunn, G. B.; Hutcheson, K. A.; Chambers, M. S.; Court, L. E.; Mohamed, A. S. R.; Fuller, C. D.; Lai, S. Y.; Rao, A. Computed Tomography Radiomics Kinetics as Early Imaging Correlates of Osteoradionecrosis in Oropharyngeal Cancer Patients. Front. Artif. Intell. 2021, 4. 10.3389/frai.2021.618469.

(30) Hardcastle, N.; Montaseri, A.; Lydon, J.; Kron, T.; Osbourne, G.; Casswell, G.; Taylor, D.; Hall, L.; McDowell, L. Dose to Medium in Head and Neck Radiotherapy: Clinical Implications for Target Volume Metrics. Phys. Imaging Radiat. Oncol. 2019, 11, 92. 10.1016/j.phro.2019.08.005.

(31) Collins, G. S.; Reitsma, J. B.; Altman, D. G.; Moons, K. Transparent Reporting of a Multivariable Prediction Model for Individual Prognosis or Diagnosis (TRIPOD): The TRIPOD Statement. BMC Med. 2015, 13 (1), 1. 10.1186/s12916-014-0241-z.

(32) International ORAL Consortium. International Expert-Based Consensus Definition, Staging Criteria, and Minimum Data Elements for Osteoradionecrosis of the Jaw: An Inter-Disciplinary Modified Delphi Study. MedRxiv Prepr. Serv. Health Sci. 2024, 2024.04.07.24305400. 10.1101/2024.04.07.24305400.

(33) Hosseinian, S.; Hemmati, M.; Dede, C.; Salzillo, T. C.; van Dijk, L. V.; Mohamed, A. S. R.; Lai, S. Y.; Schaefer, A. J.; Fuller, C. D. Cluster-Based Toxicity Estimation of Osteoradionecrosis Via Unsupervised Machine Learning: Moving Beyond Single Dose-Parameter Normal Tissue Complication Probability by Using Whole Dose-Volume Histograms for Cohort Risk Stratification. Int. J. Radiat. Oncol. Biol. Phys. 2024, 119 (5), 1569–1578. 10.1016/j.ijrobp.2024.02.021.

