## Supplementary Material for "Multi-institutional Normal Tissue Complication Probability (NTCP) Prediction Model for Mandibular Osteoradionecrosis: Results from the PREDMORN Study"

**Supplementary Material - Table of Contents**

**Supplement A.** Extract from the PREDMORN Study Protocol

**Supplement B.** Clinical and Demographic Data

**Supplement C.** Univariable Analyses

**Supplement D.** Mandible Volume Analyses

**Supplement E.** Dosimetric Data

**Supplement F.** Time to ORN Analyses

**Supplement G.** Multivariable stepwise forward logistic regression analyses

**Supplement H.** Sub-cohort analyses

**Supplement I.** Practical application of the NTCP model

**Supplement J.** TRIPOD Checklist

#### Supplement A. Extract from the PREDMORN study protocol

Table A1: Inclusion and exclusion criteria for the study as defined in the protocol [1].

| | |
|---|---|
| **Inclusion criteria** | • HNC cases treated radically with RT or chemo-radiation (CRT) and post-operative RT (PORT)+/- chemotherapy (C-PORT) using IMRT or VMAT.<br>*Specific to ORN cases:*<br>• Confirmed diagnosis of ORN (any grade). ORN defined clinically as 'an area of exposed bone in the mandible that had been present for at least 8 weeks in a previously irradiated field, in the absence of recurrent tumour' [20]. NB: cases with recurrences outside of the HN region will be included.<br>*Specific to control cases:*<br>• Any histology except for T1/2N0 Larynx cases, as these do not receive significant dose to the mandible.<br>• The selection of controls should be as per the methodology described in Figure 1. |
| **Exclusion criteria** | • Datasets without available CT volume, RT structure or RT dose DICOM files.<br>• ORN outside the mandible (e.g. maxilla). NB: if multiple ORN sites, cases will be included if at least one of the sites is the mandible.<br>• Cases with re-irradiation to the HN region.<br>• Cases treated with less than a radical dose of radiation and/or those with life expectancy at diagnosis less than 12 months. |

Table A2: Variables for data collection as proposed in the protocol [1].

| Variable | Units/format/options | Comments |
|---|---|---|
| Sex | male/female | |
| Date/year[1] of birth | dd/mm/yyyy | This is needed to calculate the age at the start of RT |
| RT start date/year | dd/mm/yyyy | |
| RT end date | dd/mm/yyyy | |
| Date/year of last follow-up | dd/mm/yyyy | This is needed to calculate the follow-up time since the end of RT |
| Primary site group | • oral cavity (OCC)<br>• oropharynx (OPC)<br>• paranasal sinus/nasopharynx (PNS/NP)<br>• larynx/hypopharynx<br>• salivary gland<br>• unknown primary | |
| TNM stage | • T: X, 0, 1, 2, 3, 4, 4a, 4b<br>• N: 0, 1, 2, 2a, 2b, 2c, 3<br>• M: 0, 1, M | Use of TNM7 notation |
| HPV | • HPV+<br>• HPV-<br>• not available | If available |
| Smoking status at start of RT | • Current: smoking at the start or within 3 months of the start of RT<br>• Previous: stopped smoking at least more than 3 months prior to the start of RT | If available |

| | | |
|---|---|---|
| | • Never: the patient has never smoked | |
| Smoking amount at start of RT | • None<br>• < 10 pack years<br>• > 10 pack years<br>• not available | If available |
| Alcohol status at start of RT | • Current: alcohol intake at the start or within 3 months of the start of RT. If available, also state amount: < 21 u/w or > 21 u/w.<br>• Previous: stopped alcohol intake at least more than 3 months prior to the start of RT<br>• Never: no alcohol intake at all | If available |
| ECOG performance status at start of RT | • 0: Able to carry normal activity without restriction<br>• 1: Restricted in physically strenous activity but ambulatory and able to carry out light work<br>• 2: Ambulatory and capable of self-care but unable to carry out any work. Up and about for more than 50% of waking hours<br>• 3: Capable only of limited self care; confined to bed or chair more than 50% of waking hours<br>• 4: Completely disabled; cannot carry out any self care; totally confined to bed or chair<br>• Not available | If available |
| Xerostomia at baseline | • G0: None<br>• G1 (Mild): Symptomatic (e.g., dry or thick saliva) without significant dietary alteration; unstimulated saliva flow >0.2ml/min<br>• G2 (Moderate): Oral intake alterations (e.g., copious water, other lubricants, diet limited to purees or soft food); unstimulated saliva flow 0.1-0.2 ml/min<br>• G3 (Severe): Inability to adequately aliment orally; tube feedings or TPN indicated, unstimulated saliva flow <0.1 ml/min<br>• not available | If available. Use the NCI CTCAE v4/v5 grading system (both versions are the same) |
| Xerostomia (grade≥2 at ≥1year post-RT) | Use NCI CTCAE v4/v5 grading system described above or the RTOG grading system for late toxicities described for ORN below. | If available. NCI CTCAE v4/v5 or RTOG |
| Pre-RT dental assessment | Yes / No | |
| Pre-RT dental extraction | Yes / No | |
| Teeth extracted | If any extractions performed, number the teeth extracted according to the FDI system | Use the FDI (World Dental Federation) numbering system |
| Date of pre-RT dental extraction | dd/mm/yyyy | This is needed to calculate the time from extraction to RT start |
| Pre-RT surgery | Primary RT (no pre-RT surgery) / Post-operative RT (PORT) | |
| Pre-RT surgery type | If available, describe the type and site of surgery | |
| RT technique | IMRT / VMAT | Cases treated with 3D conformal RT are excluded from the study |
| TPS | Specify radiotherapy treatment planning system and algorithm used to produce the clinical treatment plan (e.g. Monaco, Monte Carlo, Eclipse-Acuros XB, Eclipse-AAA, etc.) | This is needed in order to anticipate variations in exported data details between TPS |
| Dose calculation and reporting | Dm,m / Dw,m / Dw,w | This is needed to identify potential systematic biases in reported absorbed dose |
| Prescribed dose | in Gy | |
| Total number of fractions | | This is needed to calculate the fraction size |
| Treatment schedule | Please specify number of fractions per week and how these are distributed (e.g. 5 fractions/week, 1 fraction/day) | |
| Delivered dose and fractions | | If different from prescribed |
| Mandible volume | in cc | |

| | | |
|---|---|---|
| Dmean parotid glands | in Gy | |
| Dmean submandibular glands | in Gy | |
| Chemotherapy type | • None<br>• Carboplatin<br>• Cisplatin<br>• Cetuximab<br>• Other | |
| ORN | Yes / No | The info below is only relevant to ORN cases |
| Date or ORN diagnosis | dd/mm/yyyy | This is needed to calculate the time to ORN diagnosis since the end of RT |
| ORN site | Specify using the notation based upon equivalent teeth | |
| Primary ORN cause | • Spontaneous<br>• Induced - pre-RT dental extraction<br>• Induced - post-RT dental extractions<br>• Induced - dental infection<br>• Induced - dental implant<br>• Induced – denture<br>• Induced - mandible plate infection | |
| ORN grade (Notani) at diagnosis | • Stage I: ORN confined to alveolar bone<br>• Stage II: ORN limited to the alveolar bone and/or above the level of the inferior alveolar canal<br>• Stage III: ORN under the lower part of the inferior alveolar canal, with fistula or bone fracture | Use the Notani grading system |
| Peak ORN grade (Notani) | (same as above) | Use the Notani grading system |
| Date of peak grade (Notani) | dd/mm/yyyy | |
| ORN grade (CTCAE v4) at diagnosis | • Grade 1: Asymptomatic; clinical or diagnostic observations only; intervention not indicated<br>• Grade 2: Symptomatic; medical intervention indicated (e.g., topical agents); limiting instrumental ADL<br>• Grade 3: Severe symptoms; limiting self-care ADL; elective operative intervention indicated<br>• Grade 4: Life-threatening consequences; urgent intervention indicated<br>• Grade 5: Death | Use the CTCAE v4 grading system |
| Peak ORN grade (CTCAE v4) | (same as above) | Use the CTCAE v4 grading system |
| Date of peak grade (CTCAE v4) | dd/mm/yyyy | |

#### Supplement B. Clinical and demographic data

Table B1: Clinical and demographic characteristics of the train, test and external validation datasets. NA indicates missing data ('not available'); n/a indicates 'not applicable'.

| **Variables** | | **Train dataset** | | **Test dataset** | | **External dataset** | |
|---|---|---|---|---|---|---|---|
| | | **Control (N=639)** | **ORN (N=309)** | **Control (N=156)** | **ORN (N=80)** | **Control (N=39)** | **ORN (N=19)** |
| Sex (N, %) | *NA* | 0 (0.0) | 0 (0.0) | 0 (0.0) | 0 (0.0) | 0 (0.0) | 0 (0.0) |
| | *Male* | 473 (74.0) | 236 (76.4) | 125 (80.1) | 65 (81.3) | 34 (87.2) | 14 (73.7) |
| | *Female* | 166 (26.0) | 73 (23.6) | 31 (19.9) | 15 (18.8) | 5 (12.8) | 5 (26.3) |
| Age (years) | *NA (N, %)* | 0 (0.0) | 0 (0.0) | 0 (0.0) | 0 (0.0) | 0 (0.0) | 0 (0.0) |
| | *median (IQR)* | 61.0 (54.0-68.0) | 61 (55.0-66.9) | 60 (54.2-66.9) | 61 (54.0-67.0) | 61.5 (56.3-70.0) | 58.9 (55.5-64.2) |
| Follow-up time (years) | *NA (N, %)* | 0 (0.0) | 0 (0.0) | 0 (0.0) | 0 (0.0) | 0 (0.0) | 0 (0.0) |
| | *median (IQR)* | 4.4 (2.7-5.8) | 4.7 (3.2-6.1) | 4.1 (3.0-5.5) | 5.0 (3.2-6.0) | 4.8 (1.7-6.1) | 3.7 (2.4-4.7) |
| Primary Site Group (N, %) | *NA* | 5 (0.8) | 7 (2.3) | 4 (2.6) | 1 (1.3) | 0 (0.0) | 0 (0.0) |
| | *OPC* | 414 (64.8) | 185 (59.9) | 102 (65.4) | 53 (66.3) | 22 (56.4) | 10 (52.6) |
| | *OCC* | 182 (28.5) | 96 (31.1) | 41 (26.3) | 22 (27.5) | 2 (5.1) | 5 (26.3) |
| | *Larynx/hypopharynx* | 21 (3.3) | 11 (3.6) | 4 (2.6) | 2 (2.5) | 10 (25.6) | 1 (5.3) |
| | *Other* | 17 (2.7) | 10 (3.2) | 5 (3.2) | 2 (2.5) | 5 (12.8) | 3 (15.8) |
| T stage (N, %) | *NA* | 6 (0.9) | 4 (1.3) | 3 (1.9) | 2 (2.5) | 0 (0.0) | 0 (0.0) |
| | *TX* | 0 (0.0) | 0 (0.0) | 0 (0.0) | 0 (0.0) | 1 (2.6) | 1 (5.3) |
| | *T0* | 6 (0.9) | 1 (0.3) | 2 (1.3) | 0 (0.0) | 0 (0.0) | 1 (5.3) |
| | *T1* | 115 (18.0) | 44 (14.2) | 30 (19.2) | 8 (10.0) | 3 (7.7) | 5 (26.3) |
| | *T1c* | 0 (0.0) | 0 (0.0) | 0 (0.0) | 0 (0.0) | 0 (0.0) | 0 (0.0) |
| | *T2* | 201 (31.5) | 104 (33.7) | 53 (34.0) | 29 (36.3) | 17 (43.6) | 3 (15.8) |
| | *T2b* | 0 (0.0) | 0 (0.0) | 0 (0.0) | 0 (0.0) | 0 (0.0) | 0 (0.0) |
| | *T3* | 121 (18.9) | 52 (16.8) | 29 (18.6) | 14 (17.5) | 15 (38.5) | 5 (26.3) |
| | *T4* | 190 (29.7) | 104 (33.7) | 39 (25.0) | 27 (33.8) | 3 (7.7) | 4 (21.1) |
| | *T4a* | 0 (0.0) | 0 (0.0) | 0 (0.0) | 0 (0.0) | 0 (0.0) | 0 (0.0) |
| | *T4b* | 0 (0.0) | 0 (0.0) | 0 (0.0) | 0 (0.0) | 0 (0.0) | 0 (0.0) |
| N stage (N, %) | *NA* | 1 (0.2) | 1 (0.3) | 1 (0.6) | 1 (1.3) | 0 (0.0) | 0 (0.0) |
| | *N0* | 170 (26.6) | 64 (20.7) | 37 (23.7) | 21 (26.3) | 8 (20.5) | 6 (31.6) |
| | *NX* | 0 (0.0) | 0 (0.0) | 0 (0.0) | 0 (0.0) | 0 (0.0) | 0 (0.0) |
| | *N1* | 80 (12.5) | 34 (11.0) | 30 (19.2) | 14 (17.5) | 8 (20.5) | 1 (5.3) |
| | *N2* | 373 (58.4) | 199 (64.4) | 83 (53.2) | 44 (55.0) | 9 (23.1) | 6 (31.6) |
| | *N2a* | 0 (0.0) | 0 (0.0) | 0 (0.0) | 0 (0.0) | 2 (5.1) | 1 (5.3) |
| | *N2b* | 0 (0.0) | 0 (0.0) | 0 (0.0) | 0 (0.0) | 5 (12.8) | 3 (15.8) |
| | *N2c* | 0 (0.0) | 0 (0.0) | 0 (0.0) | 0 (0.0) | 5 (12.8) | 1 (5.3) |
| | *N3* | 15 (2.3) | 11 (3.6) | 5 (3.2) | 0 (0.0) | 2 (5.1) | 1 (5.3) |
| M stage (N, %) | *NA* | 253 (39.6) | 124 (40.1) | 68 (43.6) | 32 (40.0) | 0 (0.0) | 0 (0.0) |
| | *M0* | 383 (60.4) | 185 (59.9) | 88 (56.4) | 48 (60.0) | 39 (100.0) | 19 (100.0) |
| | *M1* | 3 (0.5) | 0 (0.0) | 0 (0.0) | 0 (0.0) | 0 (0.0) | 0 (0.0) |
| Mandible volume (cc) | *NA (N, %)* | 0 (0.0) | 0 (0.0) | 0 (0.0) | 0 (0.0) | 0 (0.0) | 0 (0.0) |
| | *median (IQR)* | 70.4 (56.6-84.3) | 72.5 (60.6-87.7) | 69.7 (59.0-83.5) | 74.9 (63.4-90.9) | 65.3 (56.0-72.6) | 64.1 ( 60.6-70.8) |
| HPV (N, %) | *NA* | 537 (84.0) | 267 (86.4) | 126 (80.8) | 65 (81.3) | 15 (38.5) | 7 (36.8) |
| | *HPV-* | 42 (6.6) | 21 (6.8) | 26 (16.7) | 9 (11.3) | 3 (7.7) | 4 (21.1) |
| | *HPV+* | 60 (9.4) | 21 (6.8) | 4 (2.6) | 6 (7.5) | 21 (53.8) | 8 (42.1) |
| Smoking status (N, %) | *NA* | 0 (0.0) | 0 (0.0) | 0 (0.0) | 0 (0.0) | 0 (0.0) | 0 (0.0) |
| | *Current* | 203 (31.8) | 128 (41.4) | 46 (29.5) | 37 (46.3) | 7 (17.9) | 10 (52.6) |
| | *Former* | 234 (36.6) | 107 (34.6) | 71 (45.5) | 28 (35.0) | 18 (46.2) | 5 (26.3) |
| | *Never* | 202 (31.6) | 74 (23.9) | 39 (25.0) | 15 (18.8) | 14 (35.9) | 4 (21.1) |
| Smoking amount (N, %) | *NA* | 235 (36.8) | 100 (32.4) | 49 (31.4) | 27 (33.8) | 4 (10.3) | 1 (5.3%) |
| | *> 10 pack years* | 173 (27.1) | 105 (34.0) | 56 (35.9) | 29 (36.3) | 19 (48.7) | 14 (73.7) |
| | *< 10 pack years* | 59 (9.2) | 30 (9.7) | 12 (7.7) | 9 (11.3) | 2 (5.1) | 0 (0.0) |
| | *None* | 172 (26.9) | 74 (23.9) | 39 (25.0) | 15 (18.8) | 14 (35.9) | 4 (21.1) |
| Alcohol status (N, %) | *NA* | 380 (59.5) | 193 (62.5) | 87 (55.8) | 47 (58.8) | 14 (35.9) | 0 (0.0) |
| | *Current* | 179 (28.0) | 80 (25.9) | 46 (29.5) | 29 (36.3) | 16 (41.0) | 11 (57.9) |
| | *Former* | 27 (4.2) | 12 (3.9) | 12 (7.7) | 2 (2.5) | 0 (0.0) | 0 (0.0) |
| | *Never* | 53 (8.3) | 24 (7.8) | 11 (7.1) | 2 (2.5) | 9 (23.1) | 8 (42.1) |
| Pre-RT extractions (N, %) | *NA* | 0 (0.0) | 0 (0.0) | 0 (0.0) | 0 (0.0) | 0 (0.0) | 0 (0.0) |
| | *Yes* | 270 (42.3) | 162 (52.4) | 75 (48.1) | 41 (51.3) | 15 (38.5) | 2 (10.5) |
| | *No* | 369 (57.7) | 147 (47.6) | 81 (51.9) | 39 (48.8) | 24 (61.5) | 17 (89.5) |
| Time extraction to RT (days) | *NA (N, %)* | 369 (100) | 269 (87.1) | 123 (78.8) | 70 (87.5) | 26 (66.7) | 17 (89.5) |
| | *median (IQR)* | 30.0 (20.5-49.0) | 20.0 (16.3-33.0) | 28.0 (24.0-43.0) | 26.5 (18.5-29.0) | 18.0 (15.0-32.0) | 12.5 (8.3-16.8) |

| | | | | | | | |
|---|---|---|---|---|---|---|---|
| Chemotherapy (N, %) | *NA* | 0 (0.0) | 0 (0.0) | 0 (0.0) | 0 (0.0) | 0 (0.0) | 0 (0.0) |
| | *Yes* | 402 (62.9) | 215 (69.6) | 98 (62.8) | 53 (66.3) | 36 (92.3) | 15 (78.9) |
| | *No* | 237 (37.1) | 94 (30.4) | 58 (37.2) | 27 (33.8) | 3 (7.7) | 4 (21.1) |
| RT technique (N, %) | *NA* | 0 (0.0) | 0 (0.0) | 0 (0.0) | 0 (0.0) | 0 (0.0) | 0 (0.0) |
| | *IMRT* | 478 (74.8) | 235 (76.1) | 122 (78.2) | 61 (76.3) | 3 (7.7) | 0 (0.0) |
| | *VMAT* | 161 (25.2) | 74 (23.9) | 34 (21.8)) | 19 (23.8) | 36 (92.3) | 19 (100.0) |
| Primary RT/PORT (N, %) | *NA* | 0 (0.0) | 0 (0.0) | 0 (0.0) | 0 (0.0) | 0 (0.0) | 0 (0.0) |
| | *Primary RT* | 460 (72.0) | 219 (70.9) | 117 (75.0) | 59 (73.8) | 36 (92.3) | 18 (94.7) |
| | *PORT* | 179 (28.0) | 90 (29.1) | 39 (25.0) | 21 (26.3) | 3 (7.7) | 1 (5.3) |
| Fraction size (N, %) | *NA* | 0 (0.0) | 0 (0.0) | 0 (0.0) | 0 (0.0) | 0 (0.0) | 0 (0.0) |
| | *> 2Gy/fraction* | 321 (50.2) | 164 (53.1) | 84 (53.8) | 47 (58.8) | 2 (5.1) | 2 (10.5) |
| | *<= 2Gy/fraction* | 318 (49.8) | 145 (46.9) | 72 (46.2) | 33 (41.3) | 37 (94.9) | 17 (89.5) |
| ECOG performance (N, %) | *NA* | 341 (53.4) | 166 (53.7) | 73 (46.8) | 46 (57.5) | 39 (100.0) | 13 (68.4) |
| | *0* | 139 (21.8) | 65 (21.0) | 42 (26.9) | 12 (15.0) | NA | 3 (15.8) |
| | *1* | 119 (18.6) | 55 (17.8) | 31 (19.9) | 11 (13.8) | | 2 (10.5) |
| | *2* | 38 (5.9) | 22 (7.1) | 9 (5.8) | 11 (13.8) | | 0 (0.0) |
| | *3* | 2 (0.3) | 1 (0.3) | 1 (0.6) | 0 (0.0) | | 0 (0.0) |
| | *4* | 0 (0.0) | 0 (0.0) | 0 (0.0) | 0 (0.0) | | 1 (5.3) |
| | *5* | 0 (0.0) | 0 (0.0) | 0 (0.0) | 0 (0.0) | | 0 (0.0) |
| Xerostomia baseline (N, %) | *NA* | 526 (82.3) | 251 (81.2) | 122 (78.2) | 65 (81.3) | 1 (2.6) | 5 (26.3) |
| | *0* | 99 (15.5) | 54 (17.5) | 30 (19.2) | 14 (17.5) | 29 (74.4) | 11 (57.9) |
| | *1* | 14 (2.2) | 4 (1.3) | 4 (2.6) | 1 (1.3) | 9 (23.1) | 3 (15.8) |
| | *2* | 0 (0.0) | 0 (0.0) | 0 (0.0) | 0 (0.0) | 0 (0.0) | 0 (0.0) |
| | *3* | 0 (0.0) | 0 (0.0) | 0 (0.0) | 0 (0.0) | 0 (0.0) | 0 (0.0) |
| | *4* | 0 (0.0) | 0 (0.0) | 0 (0.0) | 0 (0.0) | 0 (0.0) | 0 (0.0) |
| Xerostomia 1-year post-RT (N, %) | *NA* | 414 (64.8) | 196 (63.4) | 99 (63.5) | 52 (65.0) | 25 (64.1) | 11 (57.9) |
| | *0* | 17 (2.7) | 10 (3.2) | 5 (3.2) | 5 (6.3) | 4 (10.3) | 2 (10.5) |
| | *1* | 208 (32.6) | 103 (33.3) | 52 (33.3) | 23 (28.8) | 10 (25.6) | 6 (31.6) |
| | *2* | 0 (0.0) | 0 (0.0) | 0 (0.0) | 0 (0.0) | 0 (0.0) | 0 (0.0) |
| | *3* | 0 (0.0) | 0 (0.0) | 0 (0.0) | 0 (0.0) | 0 (0.0) | 0 (0.0) |
| | *4* | 0 (0.0) | 0 (0.0) | 0 (0.0) | 0 (0.0) | 0 (0.0) | 0 (0.0) |
| Dmean parotids (Gy) | *NA (N, %)* | 512 (80.1) | 266 (86.1) | 127 (81.4) | 64 (80.0) | 0 (0.0) | 0 (0.0) |
| | *median (IQR)* | 24.5 (16.8-32.9) | 24.2 (21.1-33.6) | 26.1 (18.6-32.5) | 22.0 (15.7-30.4) | 29.1 (26.1-33.9) | 33.3 (28.0-40.0) |
| Time to ORN (years) | *NA (N, %)* | n/a | 8 (2.6) | n/a | 3 (3.8) | n/a | 0 (0.0) |
| | *median (IQR)* | | 1.2 (0.6-2.4) | | 1.1 (0.5-1.8) | | 0.9 (0.6-1.7) |
| ORN grade Notani (N, %) | *NA* | n/a | 156 (50.5) | n/a | 41 (51.3) | n/a | 0 (0.0) |
| | *1* | | 61 (19.7) | | 11 (13.8) | | 4 (21.1) |
| | *2* | | 38 (12.3) | | 11 (13.8) | | 12 (63.2) |
| | *3* | | 54 (17.5) | | 17 (21.3) | | 3 (15.8) |
| ORN grade CTCAE (N, %) | *NA* | n/a | 195 (63.1) | n/a | 48 (60.0) | n/a | NA |
| | *1* | | 12 (3.9) | | 3 (3.8) | | |
| | *2* | | 48 (15.5) | | 10 (12.5) | | |
| | *3* | | 54 (17.5) | | 18 (22.5) | | |
| | *4* | | 0 (0.0) | | 1 (1.3) | | |
| | *5* | | 0 (0.0) | | 0 (0.0) | | |
| ORN grade Tsai (N, %) | *NA* | n/a | 187 (60.5) | n/a | 48 (60.0) | n/a | NA |
| | *1* | | 17 (5.5) | | 4 (5.0) | | |
| | *2* | | 22 (7.1) | | 11 (13.8) | | |
| | *3* | | 39 (12.6) | | 8 (10.0) | | |
| | *4* | | 44 (14.2) | | 9 (11.3) | | |

Table B2: Clinical and demographic characteristics by participating institution. NA indicates missing data ('not available'); n/a indicates 'not applicable'.

| Variables | | Institution A | | Institution B | | Institution C | | Institution D | | Institution E | | Institution F | | Institution G | |
|---|---|---|---|---|---|---|---|---|---|---|---|---|---|---|---|
| | | **Control (N=291)** | **ORN (N=157)** | **Control (N=184)** | **ORN (N=92)** | **Control (N=124)** | **ORN (N=42)** | **Control (N=90)** | **ORN (N=45)** | **Control (N=86)** | **ORN (N=43)** | **Control (N=34)** | **ORN (N=17)** | **Control (N=39)** | **ORN (N=19)** |
| Sex (N, %) | *Male* | 243 (83.5) | 136 (86.6) | 144 (78.3) | 66 (71.7) | 85 (68.5) | 25 (59.5) | 60 (66.7) | 33 (73.3) | 54 (62.8) | 29 (67.4) | 23 (67.6) | 16 (94.1) | 34 (87.2) | 14 (73.7) |
| | *Female* | 48 (16.5) | 21 (13.4) | 40 (21.7) | 26 (28.3) | 39 (31.5) | 17 (40.5) | 30 (33.3) | 12 (26.7) | 32 (37.2) | 14 (32.6) | 11 (32.4) | 1 (5.9) | 5 (12.8) | 5 (26.3) |
| Age (years) | *NA (N, %)* | 0 (0.0) | 0 (0.0) | 0 (0.0) | 0 (0.0) | 0 (0.0) | 0 (0.0) | 0 (0.0) | 0 (0.0) | 0 (0.0) | 0 (0.0) | 0 (0.0) | 0 (0.0) | 0 (0.0) | 0 (0.0) |
| | *median (IQR)* | 59.0 (54.0-66.0) | 60.0 (55.0-66.0) | 60.9 (53.3-67.0) | 62.0 (54.4-67.3) | 62.0 (56.0-70.0) | 65.0 (58.0-70.8) | 60.1 (54.3-66.7) | 58.8 (53.4-66.0) | 62.0 (56.0-68.8) | 60.0 (52.7-66.0) | 58.8 (56.0-67.8) | 66.7 (61.2-72.4) | 61.5 (56.3-70.0) | 58.9 (55.5-64.2) |
| Follow-up time (years) | *NA (N, %)* | 0 (0.0) | 0 (0.0) | 0 (0.0) | 0 (0.0) | 0 (0.0) | 0 (0.0) | 0 (0.0) | 1 (2.2) | 0 (0.0) | 0 (0.0) | 0 (0.0) | 0 (0.0) | 0 (0.0) | 0 (0.0) |
| | *median (IQR)* | 5.1 (3.4-7.9) | 5.4 (4.0-7.9) | 3.9 (3.1-5.1) | 4.0 (2.0-5.1) | 3.9 (2.2-5.0) | 4.3 (3.2-6.2) | 3.0 (1.2-4.9) | 4.3 (2.3-5.2) | 4.7 (2.2-6.2) | 5.1 (3.1-6.5) | 5.0 (3.6-5.6) | 4.1 (3.0-5.0) | 4.8 (1.7-6.1) | 3.7 (2.4-4.7) |
| Primary Site Group (N, %) | *NA* | 0 (0.0) | 0 (0.0) | 0 (0.0) | 0 (0.0) | 0 (0.0) | 0 (0.0) | 0 (0.0) | 0 (0.0) | 0 (0.0) | 0 (0.0) | 0 (0.0) | 0 (0.0) | 0 (0.0) | 0 (0.0) |
| | *OPC* | 234 (80.4) | 114 (72.6) | 104 (56.5) | 52 (56.5) | 68 (54.8) | 15 (35.7) | 52 (57.8) | 25 (55.6) | 47 (54.7) | 23 (53.5) | 20 (58.8) | 10 (58.8) | 22 (56.4) | 10 (52.6) |
| | *OCC* | 48 (16.5) | 34 (21.7) | 56 (30.4) | 28 (30.4) | 56 (45.2) | 27 (64.3) | 36 (40.0) | 19 (42.2) | 20 (23.3) | 10 (23.3) | 12 (35.3) | 6 (35.3) | 2 (5.1) | 2 (10.5) |
| | *Larynx/ hypopharynx* | 9 (3.1) | 5 (3.2) | 6 (3.3) | 3 (3.3) | 0 (0.0) | 0 (0.0) | 2 (2.2) | 1 (2.2) | 15 (17.4) | 8 (18.6) | 2 (5.9) | 1 (5.9) | 10 (25.6) | 1 (5.3) |
| | *Other* | 0 (0.0) | 4 (2.5) | 18 (9.8) | 9 (9.8) | 0 (0.0) | 0 (0.0) | 0 (0.0) | 0 (0.0) | 4 (4.7) | 2 (4.7) | 0 (0.0) | 0 (0.0) | 5 (12.8) | 3 (15.8) |
| T stage (N, %) | *NA* | 0 (0.0) | 0 (0.0) | 0 (0.0) | 1 (1.1) | 0 (0.0) | 0 (0.0) | 0 (0.0) | 0 (0.0) | 0 (0.0) | 0 (0.0) | 0 (0.0) | 1 (5.9) | 0 (0.0) | 0 (0.0) |
| | *Tx* | 3 (1.0) | 0 (0.0) | 1 (0.5) | 0 (0.0) | 0 (0.0) | 0 (0.0) | 0 (0.0) | 0 (0.0) | 0 (0.0) | 0 (0.0) | 0 (0.0) | 0 (0.0) | 1 (2.6) | 1 (5.3) |
| | *T0* | 4 (1.4) | 1 (0.6) | 9 (4.9) | 4 (4.3) | 0 (0.0) | 0 (0.0) | 0 (0.0) | 0 (0.0) | 0 (0.0) | 0 (0.0) | 0 (0.0) | 0 (0.0) | 0 (0.0) | 1 (5.3) |
| | *T1* | 62 (21.3) | 20 (12.7) | 23 (12.5) | 12 (13.0) | 28 (22.6) | 4 (9.5) | 23 (25.6) | 11 (24.4) | 8 (9.3) | 5 (11.6) | 2 (5.9) | 0 (0.0) | 3 (7.7) | 5 (26.3) |
| | *T1c* | 0 (0.0) | 0 (0.0) | 0 (0.0) | 0 (0.0) | 0 (0.0) | 0 (0.0) | 0 (0.0) | 0 (0.0) | 0 (0.0) | 0 (0.0) | 1 (2.9) | 0 (0.0) | 0 (0.0) | 0 (0.0) |
| | *T2* | 100 (34.4) | 51 (32.5) | 62 (33.7) | 28 (30.4) | 28 (22.6) | 21 (50.0) | 39 (43.3) | 19 (42.2) | 27 (31.4) | 12 (27.9) | 5 (14.7) | 2 (11.8) | 17 (43.6) | 3 (15.8) |
| | *T2b* | 1 (0.3) | 0 (0.0) | 0 (0.0) | 0 (0.0) | 0 (0.0) | 0 (0.0) | 0 (0.0) | 0 (0.0) | 0 (0.0) | 0 (0.0) | 0 (0.0) | 0 (0.0) | 0 (0.0) | 0 (0.0) |
| | *T3* | 76 (26.1) | 33 (21.0) | 28 (15.2) | 9 (9.8) | 15 (12.1) | 9 (21.4) | 12 (13.3) | 10 (22.2) | 12 (14.0) | 5 (11.6) | 7 (20.6) | 2 (11.8) | 15 (38.5) | 5 (26.3) |
| | *T4* | 39 (13.4) | 50 (31.8) | 4 (2.2) | 4 (4.3) | 0 (0.0) | 0 (0.0) | 16 (17.8) | 5 (11.1) | 39 (45.3) | 21 (48.8) | 6 (17.6) | 6 (35.3) | 3 (7.7) | 4 (21.1) |
| | *T4a* | 6 (2.1) | 2 (1.3) | 56 (30.4) | 30 (32.6) | 49 (39.5) | 8 (19.0) | 0 (0.0) | 0 (0.0) | 0 (0.0) | 0 (0.0) | 7 (20.6) | 3 (17.6) | 0 (0.0) | 0 (0.0) |
| | *T4b* | 0 (0.0) | 0 (0.0) | 1 (0.5) | 4 (4.3) | 4 (3.2) | 0 (0.0) | 0 (0.0) | 0 (0.0) | 0 (0.0) | 0 (0.0) | 6 (17.6) | 3 (17.6) | 0 (0.0) | 0 (0.0) |
| N stage (N, %) | *NA* | 0 (0.0) | 0 (0.0) | 2 (1.1) | 1 (1.1) | 0 (0.0) | 0 (0.0) | 0 (0.0) | 0 (0.0) | 0 (0.0) | 0 (0.0) | 0 (0.0) | 1 (5.9) | 0 (0.0) | 0 (0.0) |
| | *N0* | 39 (13.4) | 21 (13.4) | 60 (32.6) | 25 (27.2) | 54 (43.5) | 15 (35.7) | 24 (26.7) | 17 (37.8) | 21 (24.4) | 7 (16.3) | 11 (32.4) | 3 (17.6) | 8 (20.5) | 6 (31.6) |
| | *Nx* | 1 (0.3) | 0 (0.0) | 1 (0.5) | 0 (0.0) | 0 (0.0) | 0 (0.0) | 0 (0.0) | 0 (0.0) | 0 (0.0) | 0 (0.0) | 0 (0.0) | 0 (0.0) | 0 (0.0) | 0 (0.0) |
| | *N1* | 36 (12.4) | 16 (10.2) | 21 (11.4) | 8 (8.7) | 19 (15.3) | 5 (11.9) | 19 (21.1) | 10 (22.2) | 12 (14.0) | 6 (14.0) | 14 (41.2) | 3 (17.6) | 8 (20.5) | 1 (5.3) |
| | *N2* | 86 (29.6) | 63 (40.1) | 0 (0.0) | 0 (0.0) | 0 (0.0) | 0 (0.0) | 3 (3.3) | 0 (0.0) | 0 (0.0) | 1 (2.3) | 0 (0.0) | 1 (5.9) | 9 (23.1) | 6 (31.6) |
| | *N2a* | 10 (3.4) | 3 (1.9) | 15 (8.2) | 3 (3.3) | 3 (2.4) | 2 (4.8) | 2 (2.2) | 1 (2.2) | 7 (8.1) | 1 (2.3) | 1 (2.9) | 5 (29.4) | 0 (0.0) | 1 (5.3) |
| | *N2b* | 75 (25.8) | 30 (19.1) | 67 (36.4) | 38 (41.3) | 31 (25.0) | 16 (38.1) | 25 (27.8) | 11 (24.4) | 25 (29.1) | 15 (34.9) | 14 (41.2) | 3 (17.6) | 2 (5.1) | 3 (15.8) |
| | *N2c* | 37 (12.7) | 21 (13.4) | 13 (7.1) | 13 (14.1) | 14 (11.3) | 3 (7.1) | 15 (16.7) | 5 (11.1) | 18 (20.9) | 11 (25.6) | 4 (11.8) | 0 (0.0) | 5 (12.8) | 1 (5.3) |
| | *N3* | 7 (2.4) | 3 (1.9) | 5 (2.7) | 4 (4.3) | 3 (2.4) | 1 (2.4) | 2 (2.2) | 1 (2.2) | 3 (3.5) | 2 (4.7) | 0 (0.0) | 1 (5.9) | 2 (5.1) | 1 (5.3) |
| M stage (N, %) | *NA* | 291 (100.0) | 157 (100.0) | 11 (6.0) | 2 (2.2) | 21 (16.9) | 0 (0.0) | 0 (0.0) | 0 (0.0) | 0 (0.0) | 0 (0.0) | 0 (0.0) | 1 (5.9) | 0 (0.0) | 0 (0.0) |
| | *M0* | NA | NA | 172 (93.5) | 90 (97.8) | 103 (83.1) | 42 (100.0) | 88 (97.8) | 45 (100.0) | 86 (100.0) | 43 (100.0) | 34 (100.0) | 16 (94.1) | 39 (100.0) | 19 (100.0) |
| | *M1* | | | 1 (0.5) | 0 (0.0) | 0 (0.0) | 0 (0.0) | 2 (2.2) | 0 (0.0) | 0 (0.0) | 0 (0.0) | 0 (0.0) | 0 (0.0) | 0 (0.0) | 0 (0.0) |
| Mandible volume (cc) | *NA (N, %)* | 0 (0.0) | 0 (0.0) | 1 (0.5) | 2 (2.2) | 0 (0.0) | 0 (0.0) | 0 (0.0) | 1 (2.2) | 0 (0.0) | 0 (0.0) | 0 (0.0) | 0 (0.0) | 0 (0.0) | 0 (0.0) |
| | *median (IQR)* | 85.7 (75.4-97.3) | 86.5 (75.6-97.5) | 55.1 (47.9-64.6) | 62.5 (48.2-74.6) | 71.8 (61.2-80.3) | 72.4 (60.5-88.3) | 67.4 (58.2-73.0) | 70.5 (62.3-81.5) | 56.1 (47.5-62.7) | 54.9 (46.8-65.4) | 67.1 (52.8-73.0) | 66.5 (56.3-72.3) | 65.3 (56.0-72.6) | 64.1 (60.6-70.8) |

| | | | | | | | | | | | | | | | |
|---|---|---|---|---|---|---|---|---|---|---|---|---|---|---|---|
| HPV (N, %) | *NA* | 291 (100.0) | 157 (100.0) | 117 (63.6) | 65 (70.7) | 101 (81.5) | 38 (90.5) | 90 (100.0) | 45 (100.0) | 41 (47.7) | 17 (39.5) | 34 (100.0) | 17 (100.0) | 15 (38.5) | 7 (36.8) |
| | *HPV-* | | | 11 (6.0) | 7 (7.6) | 9 (7.3) | 3 (7.1) | | | 26 (30.2) | 20 (46.5) | | | 3 (7.7) | 4 (21.1) |
| | *HPV+* | | | 56 (30.4) | 20 (21.7) | 14 (11.3) | 1 (2.4) | | | 19 (22.1) | 6 (14.0) | | | 21 (53.8) | 8 (42.1) |
| Smoking status (N, %) | *NA* | 0 (0.0) | 0 (0.0) | 1 (0.5) | 0 (0.0) | 0 (0.0) | 0 (0.0) | 0 (0.0) | 0 (0.0) | 0 (0.0) | 0 (0.0) | 1 (2.9) | 0 (0.0) | 0 (0.0) | 0 (0.0) |
| | *Current* | 44 (15.1) | 27 (17.2) | 50 (27.2) | 47 (51.1) | 54 (43.5) | 22 (52.4) | 49 (54.4) | 32 (71.1) | 37 (43.0) | 29 (67.4) | 18 (52.9) | 12 (70.6) | 7 (17.9) | 10 (52.6) |
| | *Former* | 110 (37.8) | 72 (45.9) | 80 (43.5) | 26 (28.3) | 54 (43.5) | 15 (35.7) | 24 (26.7) | 8 (17.8) | 32 (37.2) | 12 (27.9) | 9 (26.5) | 3 (17.6) | 18 (46.2) | 5 (26.3) |
| | *Never* | 137 (47.1) | 58 (36.9) | 53 (28.8) | 19 (20.7) | 16 (12.9) | 5 (11.9) | 17 (18.9) | 5 (11.1) | 17 (19.8) | 2 (4.7) | 6 (17.6) | 2 (11.8) | 14 (35.9) | 4 (21.1) |
| Smoking amount (N, %) | *NA* | 0 (0.0) | 0 (0.0) | 38 (20.7) | 19 (20.7) | 119 (96.0) | 38 (90.5) | 90 (100.0) | 45 (100.0) | 13 (15.1) | 15 (34.9) | 34 (100.0) | 17 (100.0) | 4 (10.3) | 1 (5.3) |
| | *> 10 pack years* | 112 (38.5) | 69 (43.9) | 73 (39.7) | 45 (48.9) | 0 (0.0) | 0 (0.0) | | | 46 (53.5) | 22 (51.2) | | | 19 (48.7) | 14 (73.7) |
| | *< 10 pack years* | 42 (14.4) | 30 (19.1) | 19 (10.3) | 5 (5.4) | 0 (0.0) | 0 (0.0) | NA | NA | 27 (31.4) | 6 (14.0) | NA | NA | 2 (5.1) | 0 (0.0) |
| | *None* | 137 (47.1) | 58 (36.9) | 53 (28.8) | 19 (20.7) | 16 (12.9) | 5 (11.9) | | | 17 (19.8) | 2 (4.7) | | | 14 (35.9) | 4 (21.1) |
| Tobacco pack years | *NA (N, %)* | 0 (0.0) | 0 (0.0) | 184 (100.0) | 92 (100.0) | 124 (100.0) | 42 (100.0) | 90 (100.0) | 45 (100.0) | 14 (16.3) | 15 (34.9) | 34 (100.0) | 17 (100.0) | 39 (100.0) | 19 (100.0) |
| | *median (IQR)* | 1.0 (0.0-30.0) | 6.0 (0.0-30.0) | NA | NA | NA | NA | NA | NA | 25.0 (0.0-41.3) | 30.0 (16.5-40.0) | NA | NA | NA | NA |
| Alcohol status (N, %) | *NA* | 291 (100.0) | 157 (100.0) | 1 (0.5) | 0 (0.0) | 0 (0.0) | 0 (0.0) | 90 (100.0) | 45 (100.0) | 2 (2.3) | 3 (7.0) | 1 (2.9) | 0 (0.0) | 14 (35.9) | 0 (0.0) |
| | *Current* | | | 137 (74.5) | 71 (77.2) | 77 (62.1) | 29 (69.0) | | | 64 (74.4) | 35 (81.4) | 16 (47.1) | 10 (58.8) | 16 (41.0) | 11 (57.9) |
| | *Former* | NA | NA | 25 (13.6) | 8 (8.7) | 18 (14.5) | 3 (7.1) | NA | NA | 6 (7.0) | 1 (2.3) | 1 (2.9) | 3 (17.6) | 0 (0.0) | 0 (0.0) |
| | *Never* | | | 21 (11.4) | 13 (14.1) | 29 (23.4) | 10 (23.8) | | | 14 (16.3) | 4 (9.3) | 16 (47.1) | 4 (23.5) | 9 (23.1) | 8 (42.1) |
| Alcohol amount (N, %) | *NA* | 291 (100.0) | 157 (100.0) | 11 (6.0) | 8 (8.7) | 97 (78.2) | 33 (78.6) | 90 (100.0) | 45 (100.0) | 86 (100.0) | 43 (100.0) | 34 (100.0) | 17 (100.0) | 14 (35.9) | 0 (0.0) |
| | *> 21 units week* | | | 47 (25.5) | 32 (34.8) | 0 (0.0) | 0 (0.0) | | | | | | | 3 (7.7) | 4 (21.1) |
| | *< 21 units week* | NA | NA | 94 (51.1) | 31 (33.7) | 0 (0.0) | 0 (0.0) | NA | NA | NA | NA | NA | NA | 13 (33.3) | 7 (36.8) |
| | *None* | | | 32 (17.4) | 21 (22.8) | 27 (21.8) | 9 (21.4) | | | | | | | 9 (23.1) | 8 (42.1) |
| Pre-RT extractions (N, %) | *NA* | 0 (0.0) | 0 (0.0) | 11 (6.0) | 0 (0.0) | 0 (0.0) | 0 (0.0) | 0 (0.0) | 0 (0.0) | 0 (0.0) | 0 (0.0) | 0 (0.0) | 0 (0.0) | 0 (0.0) | 0 (0.0) |
| | *Yes* | 206 (70.8) | 64 (40.8) | 113 (61.4) | 55 (59.8) | 48 (38.7) | 16 (38.1) | 63 (70.0) | 36 (80.0) | 37 (43.0) | 31 (72.1) | 1 (2.9) | 2 (11.8) | 15 (38.5) | 2 (10.5) |
| | *No* | 85 (29.2) | 93 (59.2) | 60 (32.6) | 37 (40.2) | 76 (61.3) | 26 (61.9) | 27 (30.0) | 9 (20.0) | 49 (57.0) | 12 (27.9) | 33 (97.1) | 15 (88.2) | 24 (61.5) | 17 (89.5) |
| Time extraction to RT (days) | *NA (N, %)* | 291 (100.0) | 157 (100.0) | 107 (58.2) | 92 (100.0) | 76 (61.3) | 26 (61.9) | 90 (100.0) | 45 (100.0) | 49 (57.0) | 12 (27.9) | 34 (100.0) | 17 (100.0) | 26 (66.7) | 17 (89.5) |
| | *median (IQR)* | NA | NA | 38.0 (25.0-56.0) | NA | 32.0 (23.3-42.3) | 29.0 (18.3-40.3) | NA | NA | 20.0 (14.0-31.0) | 20.0 (18.0-26.5) | NA | NA | 18.0 (15.0-32.0) | 12.5 (8.3-16.8) |
| Chemotherapy (N, %) | *NA* | 0 (0.0) | 4 (2.5) | 1 (0.5) | 0 (0.0) | 0 (0.0) | 0 (0.0) | 0 (0.0) | 0 (0.0) | 0 (0.0) | 0 (0.0) | 0 (0.0) | 0 (0.0) | 0 (0.0) | 0 (0.0) |
| | *Yes* | 250 (85.9) | 138 (87.9) | 110 (59.8) | 59 (64.1) | 51 (41.1) | 14 (33.3) | 25 (27.8) | 13 (28.9) | 63 (73.3) | 39 (90.7) | 27 (79.4) | 15 (88.2) | 36 (92.3) | 15 (78.9) |
| | *No* | 41 (14.1) | 15 (9.6) | 73 (39.7) | 33 (35.9) | 73 (58.9) | 28 (66.7) | 65 (72.2) | 32 (71.1) | 23 (26.7) | 4 (9.3) | 7 (20.6) | 2 (11.8) | 3 (7.7) | 4 (21.1) |
| RT technique (N, %) | *NA* | 0 (0.0) | 0 (0.0) | 0 (0.0) | 0 (0.0) | 0 (0.0) | 0 (0.0) | 0 (0.0) | 0 (0.0) | 0 (0.0) | 0 (0.0) | 0 (0.0) | 0 (0.0) | 0 (0.0) | 0 (0.0) |
| | *IMRT* | 262 (90.0) | 143 (91.1) | 99 (53.8) | 53 (57.6) | 124 (100.0) | 42 (100.0) | 46 (51.1) | 25 (55.6) | 78 (90.7) | 40 (93.0) | 0 (0.0) | 0 (0.0) | 3 (7.7) | 0 (0.0) |
| | *VMAT* | 29 (10.0) | 14 (8.9) | 85 (46.2) | 39 (42.4) | 0 (0.0) | 0 (0.0) | 44 (48.9) | 20 (44.4) | 8 (9.3) | 3 (7.0) | 34 (100.0) | 17 (100.0) | 36 (92.3) | 19 (100.0) |
| Primary RT/PORT (N, %) | *NA* | 0 (0.0) | 0 (0.0) | 0 (0.0) | 0 (0.0) | 0 (0.0) | 0 (0.0) | 0 (0.0) | 0 (0.0) | 0 (0.0) | 0 (0.0) | 0 (0.0) | 0 (0.0) | 0 (0.0) | 0 (0.0) |
| | *Primary RT* | 258 (88.7) | 127 (80.9) | 111 (60.3) | 57 (62.0) | 68 (54.8) | 15 (35.7) | 59 (65.6) | 28 (62.2) | 65 (75.6) | 37 (86.0) | 26 (76.5) | 15 (88.2) | 36 (92.3) | 18 (94.7) |
| | *PORT* | 33 (11.3) | 30 (19.1) | 73 (39.7) | 35 (38.0) | 56 (45.2) | 27 (64.3) | 31 (34.4) | 17 (37.8) | 21 (24.4) | 6 (14.0) | 8 (23.5) | 2 (11.8) | 3 (7.7) | 1 (5.3) |
| Fraction size (N, %) | *NA* | 0 (0.0) | 0 (0.0) | 0 (0.0) | 0 (0.0) | 0 (0.0) | 0 (0.0) | 0 (0.0) | 0 (0.0) | 0 (0.0) | 0 (0.0) | 0 (0.0) | 0 (0.0) | 0 (0.0) | 0 (0.0) |
| | *median* | 2.1 | 2.1 | 2.2 | 2.2 | 2.0 | 2.0 | 2.0 | 2.0 | 2.0 | 2.0 | 2.1 | 2.1 | 2.0 | 2.0 |
| | *range* | 2.0-2.1 | 2.0-2.1 | 2.0-2.8 | 2.0-2.8 | 2.0-2.0 | 2.0-2.0 | 1.2-2.1 | 1.4-2.1 | 1.5-2.0 | 2.0-2.0 | 1.8-2.1 | 2.0-2.1 | 2.0-2.4 | 2.0-6.0 |
| | *> 2Gy/fraction* | 258 (88.7) | 127 (80.9) | 121 (65.8) | 62 (67.4) | 0 (0.0) | 0 (0.0) | 12 (13.3) | 8 (17.8) | 1 (1.2) | 0 (0.0) | 24 (70.6) | 15 (88.2) | 2 (5.1) | 2 (10.5) |
| | *<= 2Gy/fraction* | 33 (11.3) | 30 (19.1) | 63 (34.2) | 30 (32.6) | 124 (100.0) | 42 (100.0) | 78 (86.7) | 37 (82.2) | 85 (98.8) | 43 (100.0) | 10 (29.4) | 2 (11.8) | 37 (94.9) | 17 (89.5) |
| ECOG performance (N, %) | *NA* | 291 (100.0) | 157 (100.0) | 45 (24.5) | 20 (21.7) | 56 (45.2) | 27 (64.3) | 0 (0.0) | 0 (0.0) | 2 (2.3) | 1 (2.3) | 25 (73.5) | 11 (64.7) | 39 (100.0) | 13 (68.4) |
| | *0* | NA | NA | 86 (46.7) | 41 (44.6) | 18 (14.5) | 4 (9.5) | 39 (43.3) | 22 (48.9) | 40 (46.5) | 9 (20.9) | 5 (14.7) | 3 (17.6) | NA | 3 (15.8) |
| | *1* | | | 46 (25.0) | 30 (32.6) | 48 (38.7) | 11 (26.2) | 33 (36.7) | 17 (37.8) | 21 (24.4) | 6 (14.0) | 4 (11.8) | 3 (17.6) | | 2 (10.5) |

| | | | | | | | | | | | | | | | |
|---|---|---|---|---|---|---|---|---|---|---|---|---|---|---|---|
| | *2* | | | 7 (3.8) | 1 (1.1) | 2 (1.6) | 0 (0.0) | 16 (17.8) | 5 (11.1) | 22 (25.6) | 27 (62.8) | 0 (0.0) | 0 (0.0) | | 0 (0.0) |
| | *3* | | | 0 (0.0) | 0 (0.0) | 0 (0.0) | 0 (0.0) | 2 (2.2) | 1 (2.2) | 1 (1.2) | 0 (0.0) | 0 (0.0) | 0 (0.0) | | 0 (0.0) |
| | *4* | | | 0 (0.0) | 0 (0.0) | 0 (0.0) | 0 (0.0) | 0 (0.0) | 0 (0.0) | 0 (0.0) | 0 (0.0) | 0 (0.0) | 0 (0.0) | | 1 (5.3) |
| | *5* | | | 0 (0.0) | 0 (0.0) | 0 (0.0) | 0 (0.0) | 0 (0.0) | 0 (0.0) | 0 (0.0) | 0 (0.0) | 0 (0.0) | 0 (0.0) | | 0 (0.0) |
| Xerostomia baseline (N, %) | *NA* | 291 (100.0) | 157 (100.0) | 55 (29.9) | 33 (35.9) | 124 (100.0) | 42 (100.0) | 90 (100.0) | 45 (100.0) | 86 (100.0) | 43 (100.0) | 3 (8.8) | 0 (0.0) | 1 (2.6) | 5 (26.3) |
| | *0* | NA | NA | 106 (57.6) | 53 (57.6) | NA | NA | NA | NA | NA | NA | 31 (91.2) | 17 (100.0) | 29 (74.4) | 11 (57.9) |
| | *1* | | | 19 (10.3) | 5 (5.4) | | | | | | | 0 (0.0) | 0 (0.0) | 9 (23.1) | 3 (15.8) |
| | *2* | | | 0 (0.0) | 1 (1.1) | | | | | | | 0 (0.0) | 0 (0.0) | 0 (0.0) | 0 (0.0) |
| | *3* | | | 0 (0.0) | 0 (0.0) | | | | | | | 0 (0.0) | 0 (0.0) | 0 (0.0) | 0 (0.0) |
| | *4* | | | 4 (2.2) | 0 (0.0) | | | | | | | 0 (0.0) | 0 (0.0) | 0 (0.0) | 0 (0.0) |
| Xerostomia 1-year post-RT (N, %) | *NA* | 291 (100.0) | 157 (100.0) | 7 (3.8) | 8 (8.7) | 124 (100.0) | 42 (100.0) | 7 (7.8) | 3 (6.7) | 86 (100.0) | 43 (100.0) | 3 (8.8) | 1 (5.9) | 25 (64.1) | 11 (57.9) |
| | *0* | NA | NA | 1 (0.5) | 3 (3.3) | NA | NA | 21 (23.3) | 0 (0.0) | NA | NA | 0 (0.0) | 0 (0.0) | 4 (10.3) | 2 (10.5) |
| | *1* | | | 30 (16.3) | 24 (26.1) | | | 33 (36.7) | 11 (24.4) | | | 20 (58.8) | 1 (5.9) | 10 (25.6) | 6 (31.6) |
| | *2* | | | 146 (79.3) | 56 (60.9) | | | 25 (27.8) | 22 (48.9) | | | 10 (29.4) | 7 (41.2) | 0 (0.0) | 0 (0.0) |
| | *3* | | | 0 (0.0) | 1 (1.1) | | | 4 (4.4) | 9 (20.0) | | | 1 (2.9) | 6 (35.3) | 0 (0.0) | 0 (0.0) |
| | *4* | | | 0 (0.0) | 0 (0.0) | | | 0 (0.0) | 0 (0.0) | | | 0 (0.0) | 2 (11.8) | 0 (0.0) | 0 (0.0) |
| Dmean parotids (Gy) | *NA (N, %)* | 291 (100.0) | 157 (100.0) | 184 (100.0) | 92 (100.0) | 0 (0.0) | 0 (0.0) | 90 (100.0) | 45 (100.0) | 86 (100.0) | 43 (100.0) | 1 (2.9) | 0 (0.0) | 0 (0.0) | 0 (0.0) |
| | *median (IQR)* | NA | NA | NA | NA | 24.4 (15.8-34.1) | 22.8 (11.5-33.4) | NA | NA | NA | NA | 26.3 (22.1-32.3) | 24.2 (22.1-33.0) | 29.1 (26.1-33.9) | 33.3 (28.0-40.0) |
| Time to ORN (years) | *NA (N, %)* | | 0 (0.0) | | 0 (0.0) | | 0 (0.0) | | 0 (0.0) | | 11 (25.6) | | 0 (0.0) | | 0 (0.0) |
| | *median (IQR)* | n/a | 7.4 (4.2-10.0) | n/a | 1.1 (0.5-2.2) | n/a | 1.0 (0.6-2.4) | n/a | 0.8 (0.4-1.6) | n/a | 0.6 (0.3-1.3) | n/a | 1.2 (0.4-2.2) | n/a | 0.9 (0.6-1.7) |
| ORN grade Notani (N, %) | *NA* | n/a | NA | n/a | 0 (0.0) | n/a | 1 (2.4) | n/a | 0 (0.0) | n/a | NA | n/a | 0 (0.0) | n/a | 0 (0.0) |
| | *1* | | | | 37 (40.2) | | 14 (33.3) | | 19 (42.2) | | | | 3 (17.6) | | 4 (21.1) |
| | *2* | | | | 12 (13.0) | | 11 (26.2) | | 18 (40.0) | | | | 9 (52.9) | | 12 (63.2) |
| | *3* | | | | 43 (46.7) | | 16 (38.1) | | 8 (17.8) | | | | 5 (29.4) | | 3 (15.8) |
| ORN grade CTCAE (N, %) | *NA* | n/a | NA | n/a | NA | n/a | 0 (0.0) | n/a | 0 (0.0) | n/a | 0 (0.0) | n/a | 0 (0.0) | n/a | NA |
| | *1* | | | | | | 2 (4.8) | | 11 (24.4) | | 2 (4.7) | | 1 (5.9) | | |
| | *2* | | | | | | 17 (40.5) | | 22 (48.9) | | 9 (20.9) | | 10 (58.8) | | |
| | *3* | | | | | | 23 (54.8) | | 12 (26.7) | | 32 (74.4) | | 6 (35.3) | | |
| ORN grade Tsai (N, %) | *NA* | n/a | 0 (0.0) | n/a | NA | n/a | NA | n/a | NA | n/a | 0 (0.0) | n/a | NA | n/a | NA |
| | *1* | | 21 (13.4) | | | | | | | | 3 (7.0) | | | | |
| | *2* | | 33 (21.0) | | | | | | | | 18 (41.9) | | | | |
| | *3* | | 47 (29.9) | | | | | | | | 13 (30.2) | | | | |
| | *4* | | 56 (35.7) | | | | | | | | 9 (20.9) | | | | |

Table B3. Notani and Tsai ORN grading systems.

| | |
|---|---|
| **Notani ORN Grading System** | |
| **Stage I** | ORN confined to the alveolar bone, which is the part of the jawbone that supports the teeth. |
| **Stage II** | ORN extending to the mandibular bone, but without pathological fractures. |
| **Stage III** | ORN with extensive bone involvement, possibly including pathological fractures, or ORN extending beyond the mandibular body (e.g., to the ramus of the mandible or the skull base). |
| **Tsai ORN Grading System** | |
| **Stage 1** | Bone exposure or radiographic evidence of bone necrosis without symptoms or infection. |
| **Stage 2** | Bone exposure with symptoms such as pain, and possibly secondary infection, but without a pathologic fracture or fistula. |
| **Stage 3** | More severe involvement with complications such as pathological fracture, fistula, or involvement of adjacent structures (e.g., sking or muscles). |

#### Supplement C. Univariable analyses

Table C1: Univariable analysis for the entire PREDMORN cohort (1,263 HNC patients, 415 ORN cases) including data from all seven participating institutions for the generic cohort (including all tumour sites), and the two sub-cohorts considered, the OPC/advanced (T3/T4) larynx/hypopharynx cancer cases sub-cohort and OCC cases sub-cohort.

| | Generic cohort (all primary tumour sites) | | OPC/larynx/hypopharynx sub-cohort | | OCC sub-cohort | |
|---|---|---|---|---|---|---|
| **Variable** | **p_LRT** | **OR (95% CI)** | **p_LRT** | **OR (95% CI)** | **p_LRT** | **OR (95% CI)** |
| Sex | 0.683 | 0.94 (0.72-1.25) | 0.080 | 0.70 (0.47-1.04) | 0.845 | 1.05 (0.67-1.64) |
| Age | 0.814 | 1.00 (0.99, 1.01) | 0.770 | 1.00 (0.99-1.02) | 0.798 | 1.00 (0.98-1.02) |
| Mandible volume | 0.013 | 1.01 (1.00, 1.01) | 0.079 | 1.01 (1.00-1.02) | 0.024 | 1.01 (1.00-1.02) |
| Smoking status | 0.002 | 1.62 (1.26, 2.08) | 0.003 | 1.61 (1.18-2.20) | 0.004 | 1.95 (1.24-3.06) |
| Pack Years | 0.094 | 1.01 (1.00, 1.01) | 0.475 | 1.21 (0.72-2.04) | 0.091 | 2.13 (0.89-5.10) |
| Xerostomia 1 year | 0.015 | 0.61 (0.41, 0.91) | 0.132 | 0.67 (0.40-1.13) | 0.202 | 0.64 (0.32-1.27) |
| Pre-RT extraction | 0.026 | 1.31 (1.03, 1.66) | 0.003 | 1.58 (1.17-2.14) | 0.550 | 0.88 (0.57-1.35) |
| Time extraction to RT | 0.002 | 0.97 (0.95, 0.99) | 0.090 | 2.34 (0.88-6.23) | 0.203 | 2.64 (0.59-11.75) |
| Chemotherapy | 0.042 | 1.31 (1.01, 1.69) | 0.120 | 1.36 (0.92-2.01) | 0.072 | 1.51 (0.96-2.37) |
| PORT/PrimaryRT | 0.504 | 1.09 (0.84, 1.42) | 0.425 | 0.68 (0.27-1.74) | 0.826 | 1.06 (0.62-1.83) |
| RT technique | 0.753 | 1.04 (0.80, 1.36) | 0.643 | 1.09 (0.76-1.55) | 0.947 | 1.02 (0.64-1.61) |
| Fraction size | 0.402 | 1.11 (0.87, 1.40) | 0.072 | 1.35 (0.97-1.87) | 0.799 | 1.08 (0.60-1.92) |
| V05Gy | 0.310 | 1.01 (0.99, 1.02) | 0.173 | 1.02 (0.99-1.06) | 0.406 | 1.01 (0.99-1.03) |
| V10Gy | 0.291 | 1.01 (0.99, 1.02) | 0.084 | 1.02 (1.00-1.04) | 0.553 | 1.01 (0.99-1.02) |
| V15Gy | 0.055 | 1.01 (1.00, 1.02) | 0.004 | 1.03 (1.01-1.05) | 0.449 | 1.01 (0.99-1.02) |
| V20Gy | 0.001 | 1.01 (1.01, 1.02) | <0.001 | 1.04 (1.02-1.05) | 0.256 | 1.01 (0.99-1.02) |
| V25Gy | <0.001 | 1.02 (1.01, 1.02) | <0.001 | 1.03 (1.02-1.05) | 0.201 | 1.01 (1.00-1.02) |
| V30Gy | <0.001 | 1.02 (1.01, 1.03) | <0.001 | 1.03 (1.02-1.04) | 0.080 | 1.01 (1.00-1.02) |
| V35Gy | <0.001 | 1.02 (1.01, 1.03) | <0.001 | 1.03 (1.02-1.03) | 0.034 | 1.01 (1.00-1.02) |
| V40Gy | <0.001 | 1.02 (1.01, 1.03) | <0.001 | 1.03 (1.02-1.03) | 0.023 | 1.01 (1.00-1.02) |
| V45Gy | <0.001 | 1.02 (1.02, 1.03) | <0.001 | 1.03 (1.02-1.04) | 0.011 | 1.01 (1.00-1.03) |
| V50Gy | <0.001 | 1.02 (1.02, 1.03) | <0.001 | 1.03 (1.02-1.04) | 0.006 | 1.01 (1.00-1.03) |
| V55Gy | <0.001 | 1.03 (1.02, 1.03) | <0.001 | 1.03 (1.02-1.04) | <0.001 | 1.02 (1.01-1.03) |
| V60Gy | <0.001 | 1.03 (1.02, 1.04) | <0.001 | 1.04 (1.03-1.05) | <0.001 | 1.02 (1.01-1.03) |
| V65Gy | <0.001 | 1.03 (1.02, 1.04) | <0.001 | 1.04 (1.03-1.06) | 0.062 | 1.01 (1.00-1.03) |
| V70Gy | <0.001 | 1.05 (1.03, 1.07) | <0.001 | 1.10 (1.07-1.13) | 0.257 | 1.01 (1.00-1.03) |
| D02% | <0.001 | 1.05 (1.03, 1.07) | <0.001 | 1.06 (1.03-1.10) | 0.011 | 1.05 (1.01-1.10) |
| D05% | <0.001 | 1.06 (1.04, 1.08) | <0.001 | 1.08 (1.04-1.11) | 0.003 | 1.06 (1.02-1.11) |
| D10% | <0.001 | 1.07 (1.05, 1.09) | <0.001 | 1.08 (1.05-1.11) | 0.001 | 1.07 (1.03-1.11) |
| D15% | <0.001 | 1.07 (1.05, 1.09) | <0.001 | 1.08 (1.05-1.11) | 0.001 | 1.07 (1.03-1.11) |
| D20% | <0.001 | 1.07 (1.05, 1.09) | <0.001 | 1.08 (1.06-1.11) | <0.001 | 1.07 (1.03-1.11) |
| D25% | <0.001 | 1.071.05, 1.08) | <0.001 | 1.08 (1.05-1.10) | <0.001 | 1.06 (1.03-1.10) |
| D30% | <0.001 | 1.06 (1.05, 1.08) | <0.001 | 1.07 (1.05-1.09) | <0.001 | 1.06 (1.02-1.09) |
| D35% | <0.001 | 1.05 (1.04, 1.06) | <0.001 | 1.06 (1.05-1.08) | <0.001 | 1.05 (1.02-1.08) |
| D40% | <0.001 | 1.05 (1.04, 1.06) | <0.001 | 1.06 (1.04-1.08) | <0.001 | 1.05 (1.02-1.07) |
| D45% | <0.001 | 1.05 (1.04, 1.06) | <0.001 | 1.06 (1.04-1.08) | <0.001 | 1.04 (1.02-1.06) |
| D50% | <0.001 | 1.04 (1.03, 1.05) | <0.001 | 1.06 (1.04-1.07) | <0.001 | 1.03 (1.01-1.05) |
| D55% | <0.001 | 1.04 (1.03, 1.05) | <0.001 | 1.05 (1.05-1.07) | 0.003 | 1.02 (1.01-1.04) |
| D60% | <0.001 | 1.03 (1.02, 1.04) | <0.001 | 1.05 (1.04-1.07) | 0.021 | 1.02 (1.00-1.03) |
| D65% | <0.001 | 1.03 (1.02, 1.04) | <0.001 | 1.05 (1.03-1.07) | 0.095 | 1.01 (1.00-1.03) |
| D70% | <0.001 | 1.03 (1.02, 1.04) | <0.001 | 1.05 (1.03-1.06) | 0.219 | 1.01 (1.00-1.02) |
| D75% | <0.001 | 1.02 (1.01, 1.03) | <0.001 | 1.05 (1.03-1.06) | 0.501 | 1.00 (0.99-1.02) |
| D80% | <0.001 | 1.02 (1.01, 1.03) | <0.001 | 1.05 (1.03-1.07) | 0.869 | 1.00 (0.99-1.01) |
| D85% | <0.001 | 1.02 (1.01, 1.03) | <0.001 | 1.04 (1.03-1.06) | 0.959 | 1.00 (0.99-1.01) |
| D90% | 0.006 | 1.01 (1.00, 1.03) | <0.001 | 1.04 (1.02-1.05) | 0.814 | 1.00 (0.98-1.01) |
| D95% | 0.016 | 1.02 (1.00, 1.03) | <0.001 | 1.04 (1.02-1.06) | 0.807 | 1.00 (0.97-1.02) |
| D98% | 0.040 | 1.02 (1.00, 1.03) | 0.001 | 1.04 (1.02-1.06) | 0.837 | 1.00 (0.96-1.03) |

#### Supplement D. Mandible volume analyses

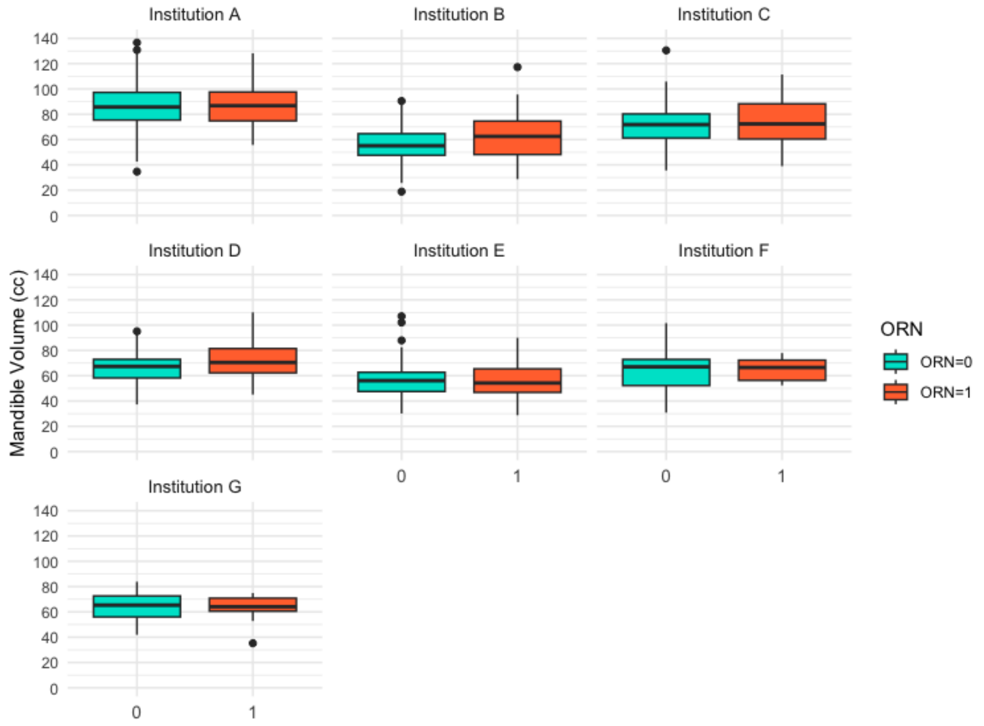


Figure D1. Institution-specific distribution of mandible volumes by group (ORN vs. control).

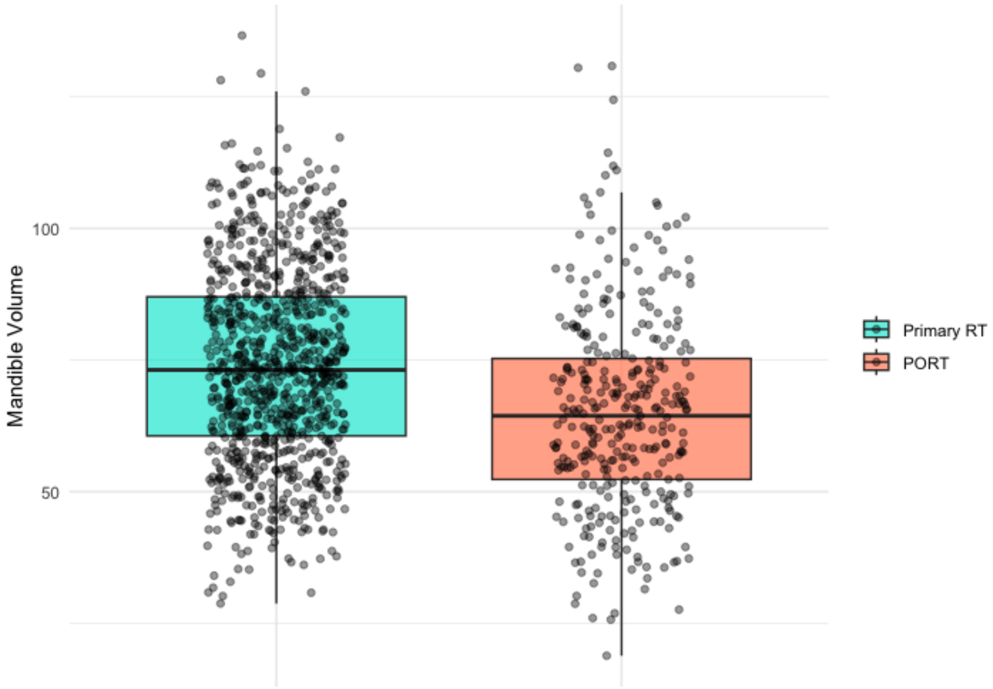


Figure D2. Distribution of mandible volume values by PORT status. Smaller mandible volumes observed in patients who underwent surgery prior RT (i.e., PORT positive status) (median 64.4 cc, IQR 52.3-75.3) compared to patients receiving RT as their primary treatment (73.1 cc, 60.6-87.0)

### Supplement E. Dosimetric data

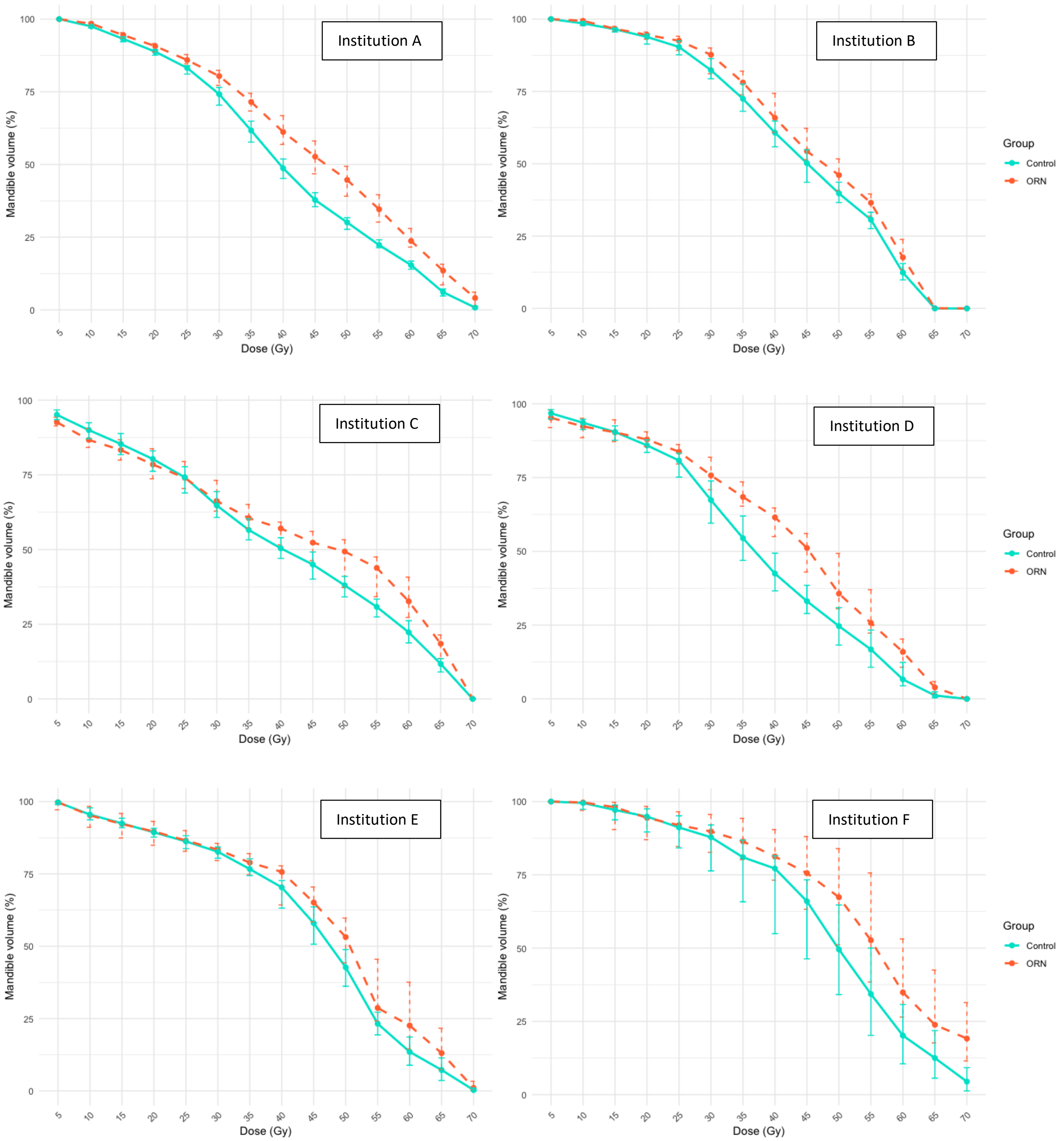

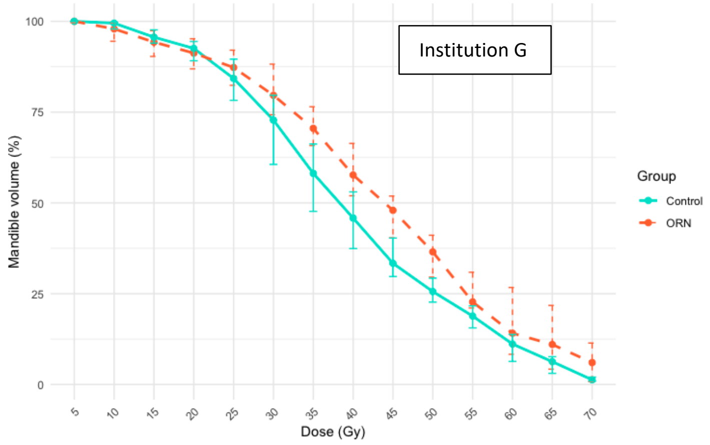


Figure E1. Institution-specific (institutions A to G) median cumulative dose-volume histogram (DVH) plots for the ORN and control groups. The medians were calculated for each DVH metric across the ORN and control groups. Error bars correspond to the 95% confidence intervals of the medians estimated using the binomial method, which identifies the lower and upper ranks corresponding to the 95% confidence bounds based on the size and distribution of the data.

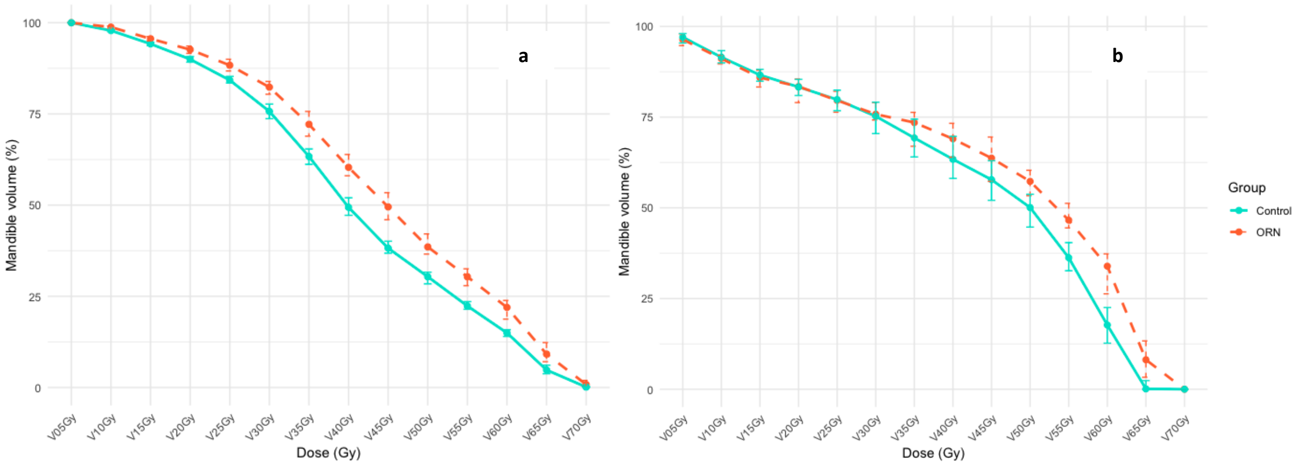


Figure E2. Median cumulative DVH plots for a) the OPC/advanced larynx/hypopharynx cancer subset and b) the OCC subset for the ORN and control groups. The medians were calculated for each DVH metric across the ORN and control groups. Error bars correspond to the 95% confidence intervals of the medians estimated using the binomial method, which identifies the lower and upper ranks corresponding to the 95% confidence bounds based on the size and distribution of the data.

#### Supplement F. Time to ORN analyses

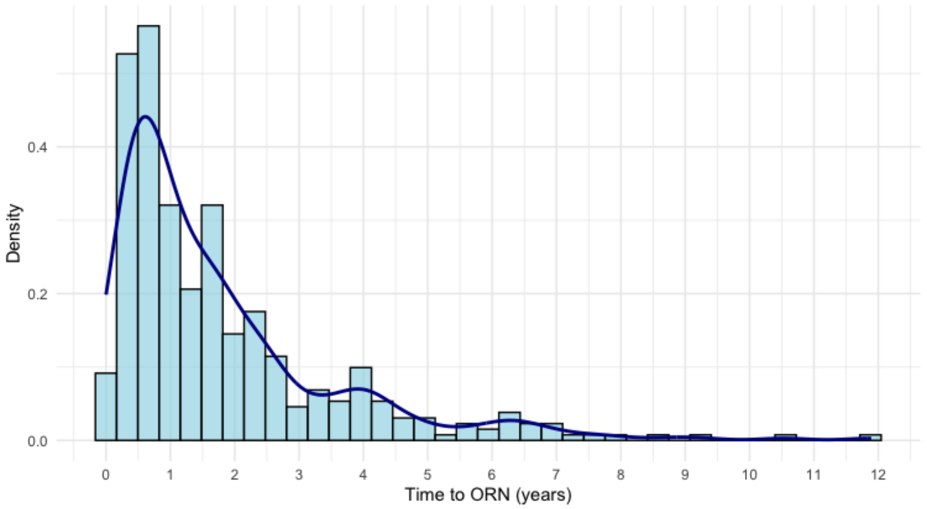


Figure F1. Distribution of time to ORN across the entire PREDMORN cohort. A density curve is overlaid to visualise the overall shape and concentration of the data, indicating the probability distribution of time to ORN.

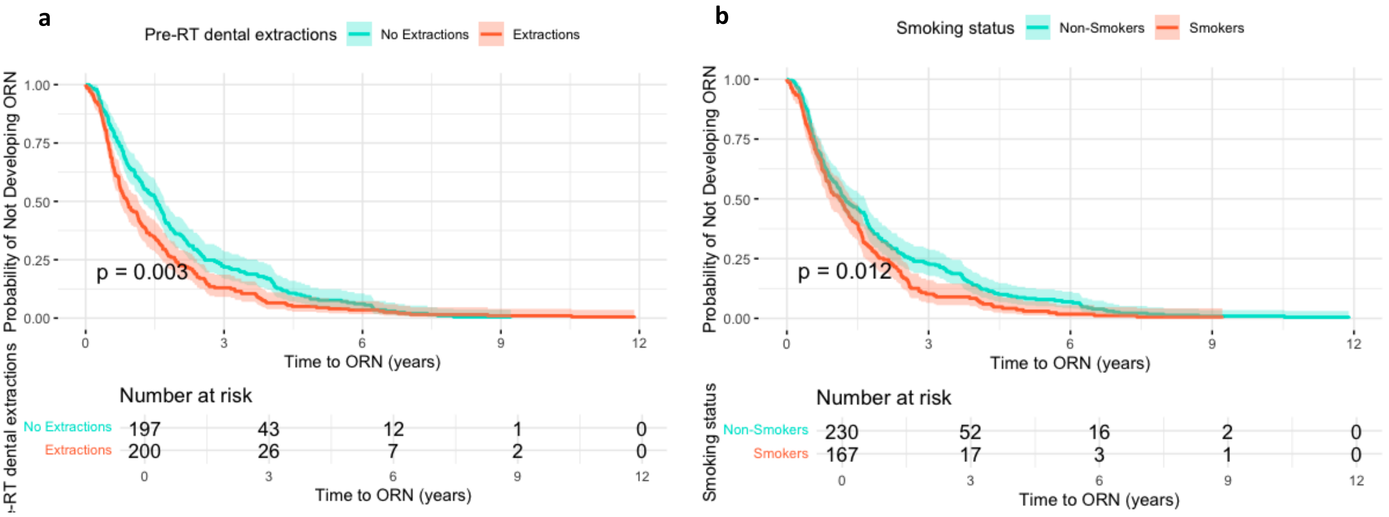


Figure F2. Kaplan-Meier survival curves, where survival corresponds to ORN-free time, stratified by sub-groups of ORN cases a) with and without pre-RT dental extractions and b) with positive (current) and negative (previous or never) smoking status. The shaded areas around each curve represent the 95% confidence intervals for the estimated survival probabilities over time. The statistical significance of the difference between the survival curves of the two groups was tested using the log-rank test, with a significance level of 0.05. The risk table below the plot displays the number of patients at risk in each group at various time points.

#### Supplement G. Multivariable stepwise forward logistic regression analyses

The Akaike Information Criterion (AIC) and the Likelihood Ratio Test (LRT) were used as the selection criteria with a threshold p-value of 0.05 for the latter. The AIC provides a means for model comparison by balancing a model's goodness of fit with model complexity (number of parameters used in the model), i.e., model parsimony. The LRT compares the goodness of fit between two nested models: a simpler model and a more complex model (i.e., simpler model with an additional variable in the forward stepwise approach).

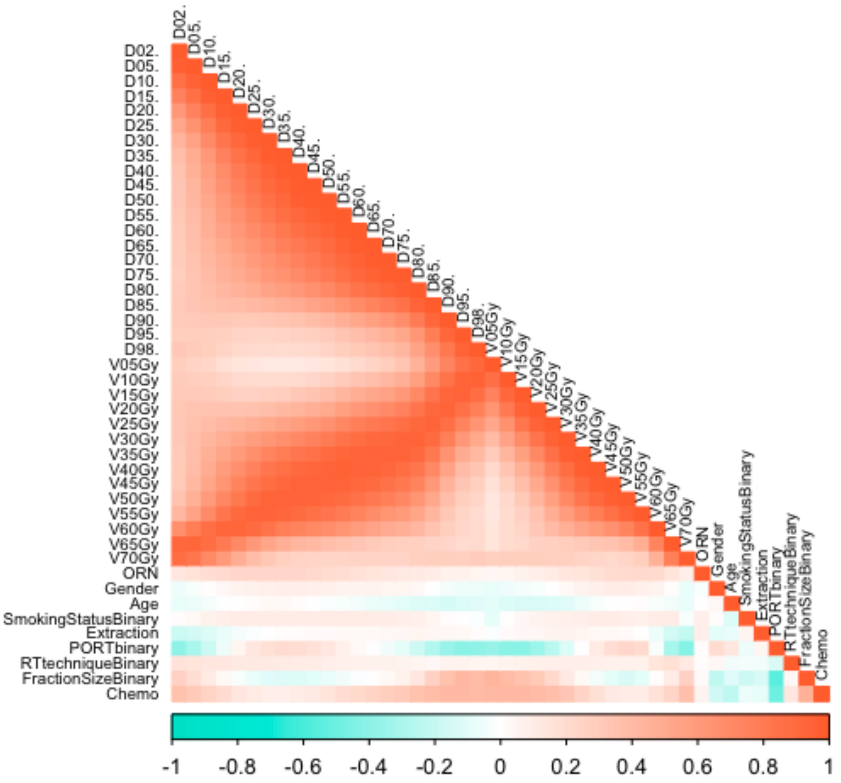


Figure G1. Spearman correlation matrix heatmap.

Table G1: Pre-selected variables and corresponding univariable p-values for the full cohort (i.e., including all primary tumour sites).

| Variable | p-value | OR (95% CI) |
|---|---|---|
| D30% | <0.0001 | 1.06 (1.04, 1.08) |
| D10% | <0.0001 | 1.07 (1.04, 1.09) |
| D60% | <0.0001 | 1.03 (1.02, 1.04) |
| V70 Gy | <0.0001 | 1.05 (1.03, 1.07) |
| V25 Gy | 0.0001 | 1.02 (1.01, 1.03) |
| Pre-RT dental extraction | 0.003 | 1.51 (1.15, 1.98) |
| Smoking status | 0.004 | 1.52 (1.15, 2.01) |
| D95% | 0.006 | 1.02 (1.01, 1.04) |
| Chemotherapy | 0.044 | 1.35 (1.01, 1.80) |
| Fraction size | 0.412 | 1.12 (0.85, 1.47) |
| Sex | 0.434 | 0.88 (0.64, 1.21) |
| RT technique | 0.677 | 1.07 (0.78, 1.47) |
| PORT | 0.722 | 1.06 (0.78, 1.43) |
| Age | 0.977 | 1.00 (0.99, 1.01) |

Table G2: Models evaluated during the stepwise forward selection process (all primary tumour sites). The thick line box indicates the model that was selected for this cohort.

| Step | Variables | β | OR | p_val | AIC | BIC | p_LRT |
|---|---|---|---|---|---|---|---|
| 1 | Intercept | -3.849 | 0.02 | | 1144.0 | 1153.7 | <0.0001 |
| | D30% | 0.058 | 1.06 | <0.0001 | | | |
| 2 | Intercept | -4.102 | 0.02 | | 1136.7 | 1151.2 | 0.002 |
| | D30% | 0.059 | 1.06 | <0.0001 | | | |
| | Dental extraction | 0.437 | 1.55 | 0.002 | | | |
| 3 | Intercept | -3.728 | 0.02 | | 1133.5 | 1152.9 | 0.023 |
| | D30% | 0.050 | 1.05 | <0.0001 | | | |
| | Extraction | 0.492 | 1.64 | 0.001 | | | |
| | V70Gy | 0.024 | 1.02 | 0.030 | | | |
| 4 | Intercept | -3.749 | 0.02 | | 1154.8 | 1154.8 | 0.026 |
| | D30% | 0.048 | 1.05 | <0.0001 | | | |
| | Dental extraction | 0.481 | 1.62 | 0.001 | | | |
| | V70Gy | 0.025 | 1.03 | 0.024 | | | |
| | Smoking status | 0.333 | 1.39 | 0.026 | | | |
| 5 | Intercept | -4.029 | 0.02 | | 1159.5 | 1159.5 | 0.140 |
| | D30% | 0.051 | 1.05 | <0.0001 | | | |
| | Dental extraction | 0.499 | 1.65 | 0.001 | | | |
| | V70Gy | 0.020 | 1.02 | 0.084 | | | |
| | Smoking status | 0.370 | 1.45 | 0.015 | | | |
| | Fraction Size | 0.231 | 1.26 | 0.140 | | | |

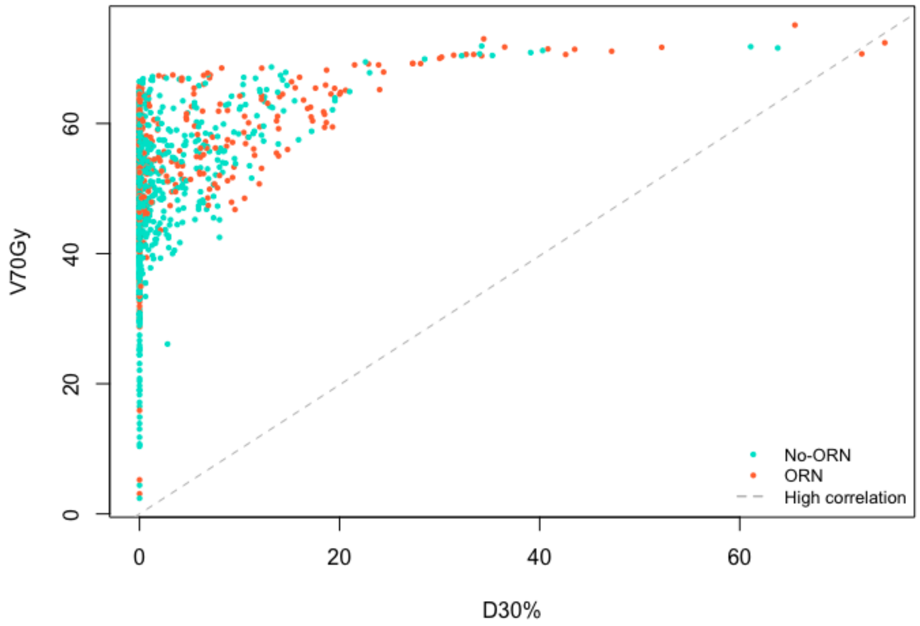


Figure G2. Scatter plot demonstrating the low collinearity between the two dosimetric variables selected in the final NTCP model, $D_{30\%}$ and $V_{70Gy}$.

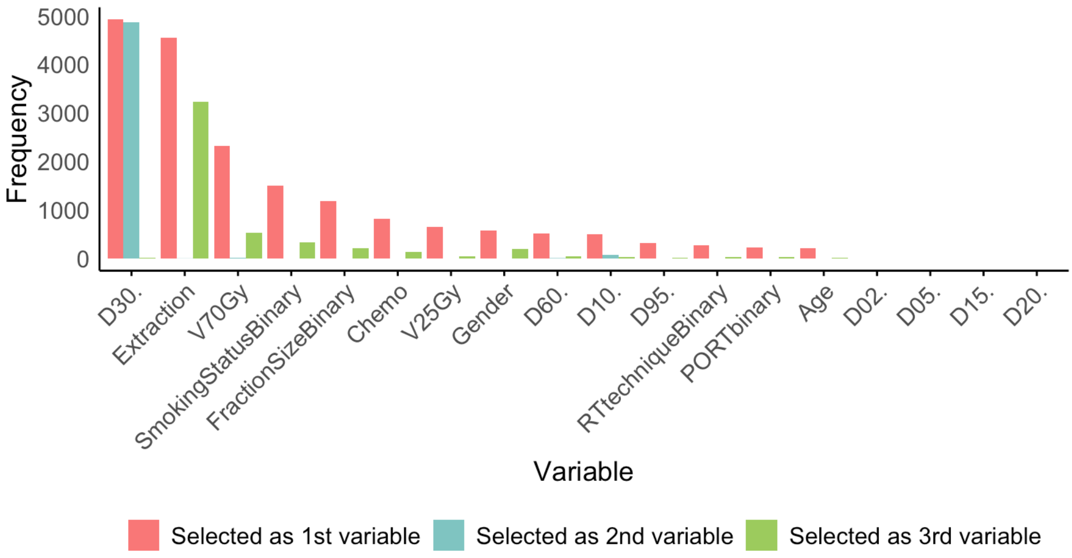


Figure G3: Bootstrapped variable selection frequency results.

#### Supplement H. Sub-cohort analyses

Table H1. Model performance results of the ORN NTCP model on the sub-cohorts of patients treated with OPC/advanced larynx/hypopharynx cancer or OCC. The confidence interval (CI) for the area under the ROC curve (AUC) has been calculated using the DeLong method.

| **ORN NTCP model performance on the OPC/advanced larynx/hypopharynx cancer sub-cohort** | | |
|---|---|---|
| | **Test (N=156)** | **External validation (N=32)** |
| ROC AUC (95% CI) | 0.75 (0.67-0.83) | 0.75 (0.58-0.92) |
| Nagelkerke $R^2$ | 0.253 | 0.255 |
| Brier score | 0.186 | 0.167 |
| Log Loss | 0.544 | 0.497 |
| **ORN NTCP model performance on the OCC sub-cohort** | | |
| | **Test (N=68)** | **External validation (N=7)** |
| ROC AUC (95% CI) | 0.53 (0.38-0.68) | n/a* |
| Nagelkerke $R^2$ | 0.008 | |
| Brier score | 0.212 | |
| Log Loss | 0.615 | |

* External validation of the model on the OCC sub-cohort was not attempted due to the small sample sizes for this data subset (N=7).

Table H2. Model performance results of the ORN NTCP model on the sub-cohorts of patients treated with Primary RT or PORT. The confidence interval (CI) for the area under the ROC curve (AUC) has been calculated using the DeLong method.

| **ORN NTCP model performance on the Primary RT sub-cohort** | | |
|---|---|---|
| | **Test (N=176)** | **External validation (N=54)** |
| ROC AUC (95% CI) | 0.70 (0.62-0.79) | 0.72 (0.58-0.86) |
| Nagelkerke $R^2$ | 0.129 | 0.209 |
| Brier score | 0.170 | 0.190 |
| Log Loss | 0.512 | 0.555 |
| **ORN NTCP model performance on the PORT sub-cohort** | | |
| | **Test (N=60)** | **External validation (N=4)** |
| ROC AUC (95% CI) | 0.60 (0.45-0.74) | n/a* |
| Nagelkerke $R^2$ | 0.065 | |
| Brier score | 0.210 | |
| Log Loss | 0.606 | |

* External validation of the model on the PORT sub-cohort was not attempted due to the small sample sizes for this data subset (N=4).

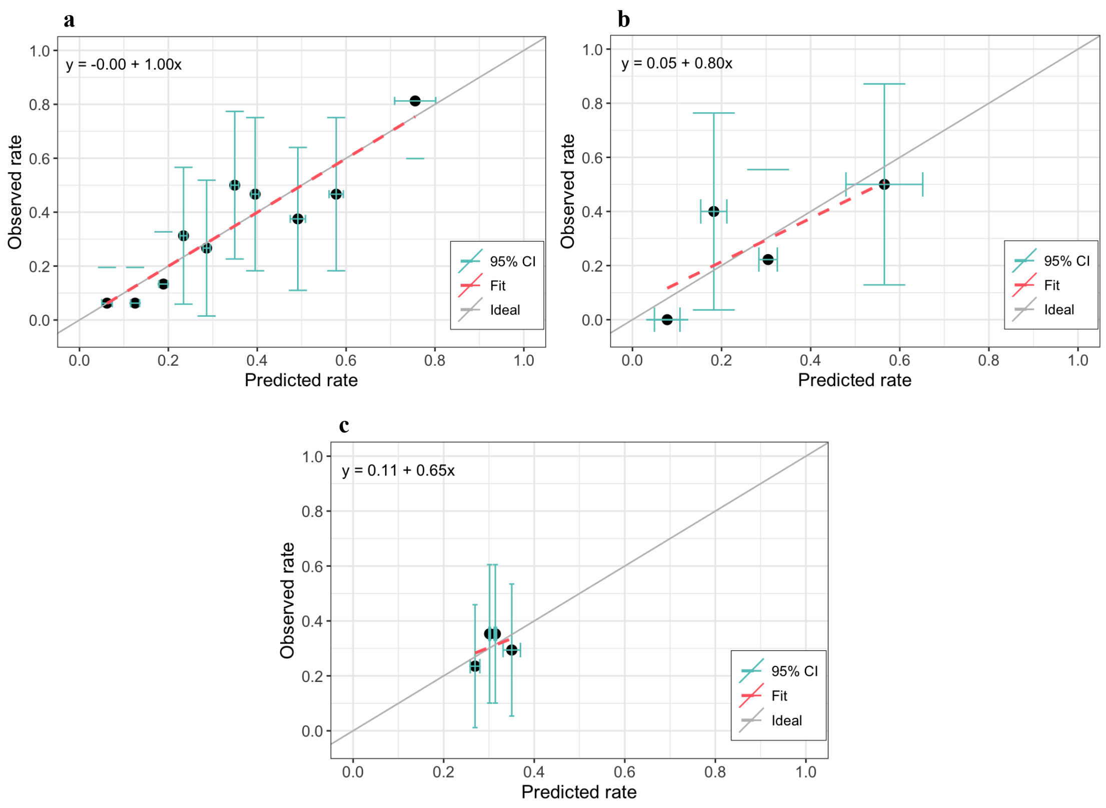


Figure H: Calibration curves when testing the NTCP model on the a) test and b) external validation datasets of the OPC/advanced larynx/hypopharynx cancer sub-cohort, and c) on the test dataset of the oral cavity cancer (OCC) sub-cohort. The number of bins in the calibration plots was adjusted in each plot based on dataset size for each cohort/sub-cohort to optimise representation of the data.

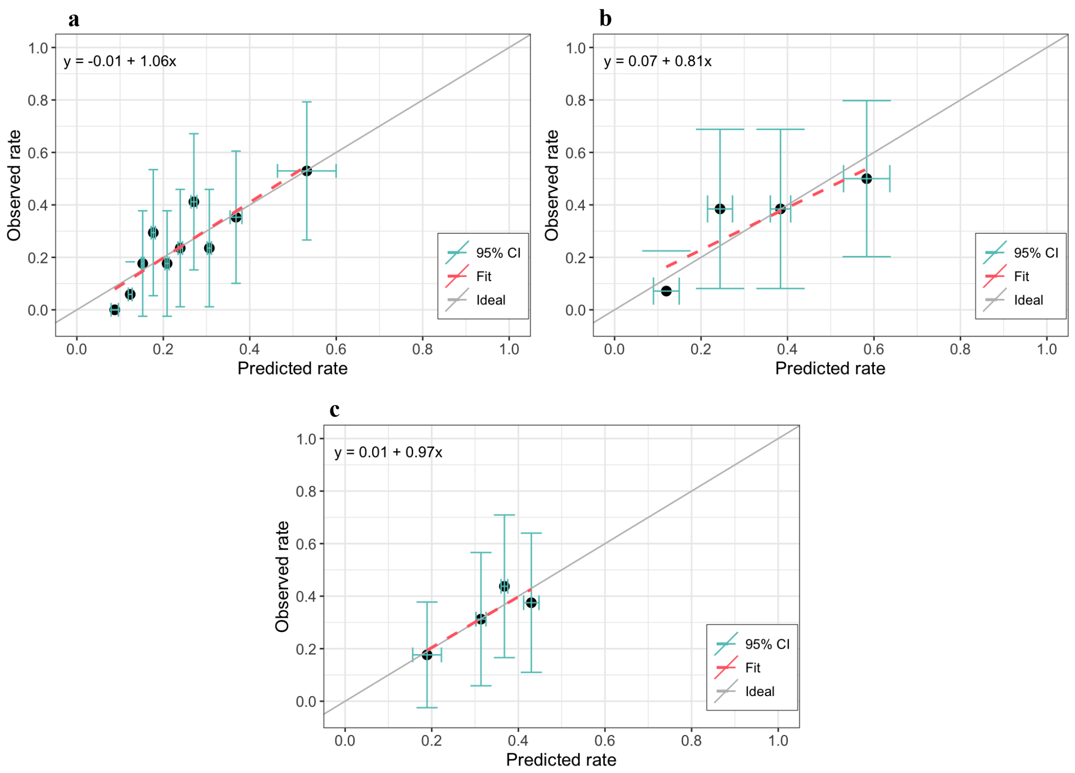


Figure H2: Calibration curves when testing the NTCP model on the a) test and b) external validation datasets of the Primary RT sub-cohort, and c) on the test dataset of the post-operative RT (PORT) sub-cohort.

#### Supplement I. Practical application of the ORN NTCP model

In our study, we used a 2:1 matched case-control approach that resulted in an artificially elevated ORN incidence rate of 32.9%. Consequently, the predicted ORN risk probabilities should be considered as relative rather than absolute. To apply our ORN NTCP model on an external dataset and obtain the adjusted ORN risk probabilities for the target population ($NTCP_{target}$), the original predicted probabilities ($NTCP_{original}$) must be adjusted based on the baseline ORN risk of the target population.

The logistic regression-based NTCP model developed on the original population can be expressed as a combination of predictors $x_i$ with their corresponding weights or model coefficients $\beta_i$:

$$NTCP_{original} = \frac{1}{(1+e^{-s})}$$

$$\text{where } s = \beta_{0,original} + \beta_1 x_1 + \cdots + \beta_i x_i$$

The baseline ORN risk for the original population is given by the intercept of the model, $\beta_{0,original}$, which corresponds to the model's implied ORN risk when all predictors are set to zero. The resulting baseline odds for the original population is given by:

$$Odds_{baseline,original} = e^{-\beta_{0,original}}$$

For the target population, the best available estimate of the baseline risk is the known ORN incidence rate ($P_{target}$) for that population and the baseline odds for the target population can be calculated as:

$$Odds_{baseline,target} = \frac{P_{target}}{1-P_{target}}$$

Finally, the model's intercept can be adjusted as follows:

$$\beta_0' = \beta_0 + ln\left(\frac{Odds_{baseline,original}}{Odds_{baseline,target}}\right)$$

And the adjusted NTCP model ($NTCP_{target}$) can be defined as:

$$NTCP_{target} = \frac{1}{(1+e^{-s'})}$$

$$\text{where } s' = \beta_0' + \beta_1 x_1 + \cdots + \beta_i x_i$$

*Worked example:*

Say we would like to apply our NTCP model on data from a patient from a new target population who has had pre-RT dental extractions, has never smoked and has been irradiated with a dose distribution that results in a mandible $D_{30\%}$ of 30 Gy and $V_{70Gy}$ of 2%.

Using the intercept and coefficients from Table 2, the predicted relative ORN risk directly calculated with our model $NTCP_{original}$ would be calculated as follows:

$$NTCP_{original} = \frac{1}{(1+e^{-s})} = \frac{1}{(1+e^{-(-1.78)})} = 0.145 = \mathbf{14.5\%}$$

$$\text{where } s = -3.749 + 0.048 \times 30 + 0.481 \times 1 + 0.025 \times 2 + 0.333 \times 0 = -1.78$$

$$\text{and } Odds_{baseline,original} = e^{-\beta_{0,original}} = e^{-(-3.749)} = 0.0235$$

However, if the baseline ORN risk for the target population that this patient belongs to is 5%, the baseline odds for the target population can be calculated as:

$$Odds_{baseline,target} = \frac{P_{target}}{1 - P_{target}} = \frac{0.05}{1 - 0.05} = 0.0526$$

The model's intercept would be adjusted as:

$$\beta_0' = \beta_0 + ln\left(\frac{Odds_{baseline,original}}{Odds_{baseline,target}}\right) = -3.749 + \ln\left(\frac{0.0235}{0.0526}\right) = -4.551$$

Finally, the adjusted ORN risk for this patient would be:

$$NTCP_{target} = \frac{1}{(1 + e^{-s'})} = \frac{1}{(1 + e^{-(-2.58)})} = 0.070 = \mathbf{7.0}\%$$

where $s' = -4.551 + 0.048 \times 30 + 0.481 \times 1 + 0.025 \times 2 + 0.333 \times 0 = -2.58$

#### Supplement J. TRIPOD checklist

| Section/Topic | | | Checklist Item | Page |
|---|---|---|---|---|
| **Title and abstract** | | | | |
| Title | 1 | D;V | Identify the study as developing and/or validating a multivariable prediction model, the target population, and the outcome to be predicted. | 1 |
| Abstract | 2 | D;V | Provide a summary of objectives, study design, setting, participants, sample size, predictors, outcome, statistical analysis, results, and conclusions. | 2 |
| **Introduction** | | | | |
| Background and objectives | 3a | D;V | Explain the medical context (including whether diagnostic or prognostic) and rationale for developing or validating the multivariable prediction model, including references to existing models. | 3 |
| | 3b | D;V | Specify the objectives, including whether the study describes the development or validation of the model or both. | 4 |
| **Methods** | | | | |
| Source of data | 4a | D;V | Describe the study design or source of data (e.g., randomized trial, cohort, or registry data), separately for the development and validation data sets, if applicable. | 4 |
| | 4b | D;V | Specify the key study dates, including start of accrual; end of accrual; and, if applicable, end of follow-up. | 4 |
| Participants | 5a | D;V | Specify key elements of the study setting (e.g., primary care, secondary care, general population) including number and location of centres. | 4 |
| | 5b | D;V | Describe eligibility criteria for participants. | 4/Sup. A |
| | 5c | D;V | Give details of treatments received, if relevant. | 11/Sup. B |
| Outcome | 6a | D;V | Clearly define the outcome that is predicted by the prediction model, including how and when assessed. | 6/Sup. B |
| | 6b | D;V | Report any actions to blind assessment of the outcome to be predicted. | |
| Predictors | 7a | D;V | Clearly define all predictors used in developing or validating the multivariable prediction model, including how and when they were measured. | 5/Sup. A, B |
| | 7b | D;V | Report any actions to blind assessment of predictors for the outcome and other predictors. | |
| Sample size | 8 | D;V | Explain how the study size was arrived at. | 5 |
| Missing data | 9 | D;V | Describe how missing data were handled (e.g., complete-case analysis, single imputation, multiple imputation) with details of any imputation method. | 8 |
| Statistical analysis methods | 10a | D | Describe how predictors were handled in the analyses. | 5 |
| | 10b | D | Specify type of model, all model-building procedures (including any predictor selection), and method for internal validation. | 6/7 |
| | 10c | V | For validation, describe how the predictions were calculated. | 6/7 |
| | 10d | D;V | Specify all measures used to assess model performance and, if relevant, to compare multiple models. | 6/7 |
| | 10e | V | Describe any model updating (e.g., recalibration) arising from the validation, if done. | Sup. I |
| Risk groups | 11 | D;V | Provide details on how risk groups were created, if done. | |
| Development vs. validation | 12 | V | For validation, identify any differences from the development data in setting, eligibility criteria, outcome, and predictors. | Sup. B, D, E |
| **Results** | | | | |
| Participants | 13a | D;V | Describe the flow of participants through the study, including the number of participants with and without the outcome and, if applicable, a summary of the follow-up time. A diagram may be helpful. | 7/8 |
| | 13b | D;V | Describe the characteristics of the participants (basic demographics, clinical features, available predictors), including the number of participants with missing data for predictors and outcome. | 11/Sup. B, D, E |
| | 13c | V | For validation, show a comparison with the development data of the distribution of important variables (demographics, predictors and outcome). | Sup. B, D, E |
| Model development | 14a | D | Specify the number of participants and outcome events in each analysis. | Sup. B, D, E |
| | 14b | D | If done, report the unadjusted association between each candidate predictor and outcome. | 8/Sup. C |
| Model specification | 15a | D | Present the full prediction model to allow predictions for individuals (i.e., all regression coefficients, and model intercept or baseline survival at a given time point). | 13/Sup. I |
| | 15b | D | Explain how to the use the prediction model. | Sup. I |
| Model performance | 16 | D;V | Report performance measures (with CIs) for the prediction model. | 14/Sup. G, H |
| Model-updating | 17 | V | If done, report the results from any model updating (i.e., model specification, model performance). | |
| **Discussion** | | | | |
| Limitations | 18 | D;V | Discuss any limitations of the study (such as nonrepresentative sample, few events per predictor, missing data). | 15-19 |
| Interpretation | 19a | V | For validation, discuss the results with reference to performance in the development data, and any other validation data. | 15-19 |
| | 19b | D;V | Give an overall interpretation of the results, considering objectives, limitations, results from similar studies, and other relevant evidence. | 15-19 |
| Implications | 20 | D;V | Discuss the potential clinical use of the model and implications for future research. | 15-19 |
| **Other information** | | | | |
| Supplementary information | 21 | D;V | Provide information about the availability of supplementary resources, such as study protocol, Web calculator, and data sets. | Supplementary material |
| Funding | 22 | D;V | Give the source of funding and the role of the funders for the present study. | 13 |
